# Early brain-wide disruption of sleep microarchitecture in Amyotrophic Lateral Sclerosis

**DOI:** 10.1101/2025.03.28.25324779

**Authors:** Christina Lang, Simon J. Guillot, Dorothee Lule, Luisa T. Balz, Antje Knehr, Patrick Weydt, Johannes Dorst, Katharina Kandler, Hans-Peter Muller, Jan Kassubek, Laura Wassermann, Sandrine Da Cruz, Francesco Roselli, Albert C. Ludolph, Matei Bolborea, Luc Dupuis

## Abstract

Amyotrophic lateral sclerosis (ALS), the major adult-onset motor neuron disease, is preceded by an early period unrelated to the motor system, including altered sleep, with increased wake and decreased NREM sleep phases. Whether these alterations in sleep macroarchitecture are associated, or even preceded by abnormalities in sleep-related EEG hallmarks is unknown. Here we used polysomnography to characterize sleep microarchitecture in the early phases of ALS. We observed a brain-wide decrease in density of sleep spindles, slow oscillations and k-complexes, three sleep-related EEG signals, in both early-stage ALS patients and presymptomatic gene carriers. These alterations in sleep spindles were correlated with cognitive performance, particularly of scores in memory, verbal fluency and speech, in both cohorts. Importantly, alterations in sleep microarchitecture were replicated in 3 mouse models and decreases in sleep spindles were rescued by MCH intracerebroventricular supplementation or oral administration of a dual orexin receptor antagonist. Thus, sleep microarchitecture is associated with cognitive deficits and causally related to aberrant MCH and orexin signaling in ALS.

## Introduction

Amyotrophic lateral sclerosis (ALS) is a fatal and rapidly progressive disease affecting upper and lower motor neurons in adults, with a median survival of three to four years after onset of motor symptoms. Onset occurs usually between 60 and 70 years of age ^1,2^ and most cases do not show a family history. However 5-10% of ALS cases are familial cases, with more than 40 distinct genes currently associated, and mutations in *C9ORF72, SOD1*, *TARDBP* and *FUS*, as major genetic causes of ALS^1,2^.

It is generally considered that ALS patients do not show clinical manifestations before the onset of symptoms. As a matter of fact, the increase in circulating neurofilament levels, that is considered a reliable biomarker of (motor) axonal injury, is observed at onset of motor symptoms but not one to two years before onset ^3,4^. Clinically, most presymptomatic gene carriers do not show even mild motor impairment 2 years before onset of motor symptoms ^4,5^. However, future ALS patients show a number of early non-motor signs, such as weight loss ^6–10^ or cognitive impairment ^11,12^, many years before onset of motor symptoms.

Recently, we identified sleep alterations as a novel early non-motor sign. We studied sleep in two cohorts of ALS patients devoid of respiratory insufficiency and presymptomatic gene carriers. In both cohorts, we observed significant defects in sleep microarchitecture characterized by increased wake and decreased deep sleep (NREM2/3)^13^, that were detectable at least 10-15 years before the anticipated onset of motor symptoms in presymptomatic gene carriers. Importantly, the severity of alterations in sleep correlated with cognitive scores^13^ and were mirrored by mouse models of familial ALS ^13^.

While this study established sleep defects as an early phenotype in future ALS patients, we did not characterize how sleep-related EEG hallmarks were affected in ALS. The different sleep states are characterized by various alterations in EEG recordings that are related to the activation of specific cortical and subcortical pathways and constitute sleep microarchitecture. These EEG hallmarks of sleep include sleep spindles, a burst of 12-15 Hz sinusoidal cycles in EEG^14^, as well as slow oscillations (<1Hz) and K-complexes, that are all involved in NREM sleep continuity and the role of sleep in memory consolidation and cognitive function^14,15^. A defect in sleep microarchitecture could be caused by specific neuroanatomical pathways, and also serve as a possible biomarker for sleep deficits. Here, we characterized sleep microarchitecture components in ALS patients, presymptomatic gene carriers and mouse models^13^. We observed a brain wide decrease in sleep spindles, slow oscillations and K-complexes, that were correlated to cognitive function. We also observed similar alterations in mouse models and showed their rescue by MCH supplementation or a dual-orexin receptor antagonist. Thus, sleep microarchitecture dynamics are early affected in ALS and relates to hypothalamic dysfunction.

## Results

### Early ALS patients exhibit brain-wide decreased sleep spindles density

To examine the extent of alterations in sleep microarchitecture in individuals with early-stage amyotrophic lateral sclerosis (ALS), we took advantage of polysomnography acquired in a previous cohort study^13^. Characteristics of included patients are provided in **Table 1**. Importantly, we prospectively excluded patients and controls with abnormal capnography, thus ruling out that the observed sleep defects were secondary to respiratory insufficiency. No significant differences were observed between the ALS patients and the control group with regard to age, sex or body mass index.

**Table 1.** Descriptive statistics of the study population of ALS patients and healthy controls. (SEM: standard error of means; BMI: body mass index; ALSFRSr: Amyotrophic Lateral Sclerosis Functional Rating Scale-Revised; ns p_value_>0.05, non-parametric Kruskal-Wallis’ test).

|  |  | ALS | Controls | p<sub>value</sub> | Reference |
| --- | --- | --- | --- | --- | --- |
| Gender | Women (%) | 14 (42.4) | 10 (31.3) |  | 13 |
|  | Men (%) | 19 (57.6) | 22 (68.7) |  |  |
| BMI Mean (±SEM) |  | 26.38 (±0.78) | 26.63 (±0.77) | 0.885 |  |
| Age Mean (±SEM) |  | 58.56 (±1.74) | 56.22 (±2.81) | 0.652 |  |
| ALSFRS<sub>r</sub> Mean (±SEM) |  | 40.51 (0.78) | — |  |  |

Consistent with the previously observed strong defect in NREM2 and 3, we observed a pronounced decrease in sleep spindle density and their root mean square in ALS patients (**Figure 1A-B**), while their amplitude was unchanged (**Figure 1B**). Topographically, we observed a significant decrease in density in both frontal and motor cortex electrodes (**Figure 1C**) while the root mean square of sleep spindles was only significantly decreased in motor cortex electrodes (C3/C4) but not in frontal cortex electrodes (F3/F4) (**Figure 1C**). Consistent with decreased sleep spindles density, we observed similar brain-wide reductions in slow oscillations and K-complexes in individuals with early-stage ALS (**Supplementary Figures 1 and 2**). Thus sleep microarchitecture is altered in early-stage ALS patients at the whole brain level.

**Figure 1:**
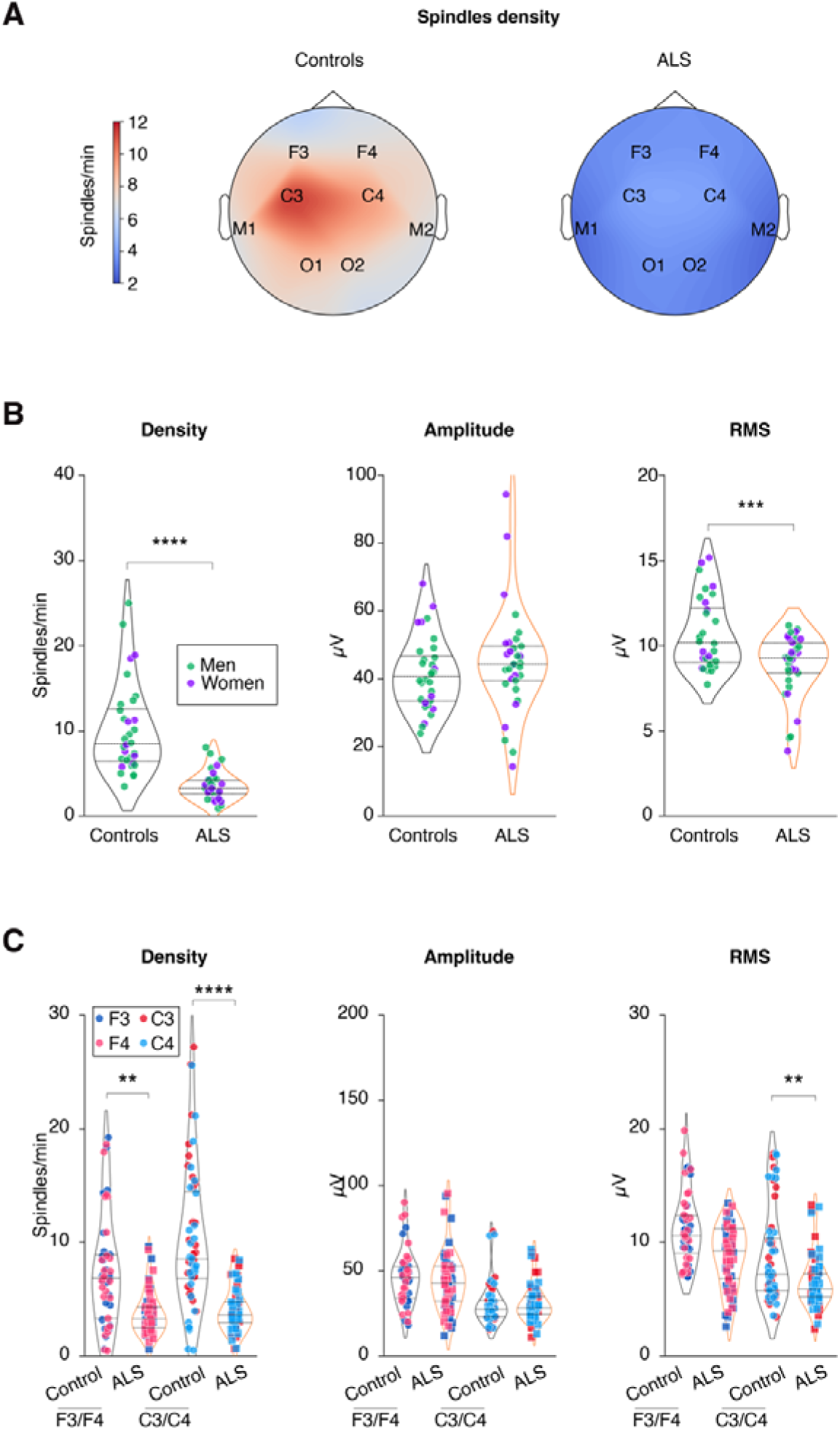
Sleep spindles alterations in early ALS patients. **(A)** Topographic map across all electrodes of sleep spindle density in controls and ALS patients. **(B)** Quantification of sleep spindle density, amplitude and root-mean-square (RMS). Men are shown in green and women in purple. **(C)** Quantification of sleep spindle density, amplitude and root-mean-square (RMS) across F3/F4 and C3/C4 electrodes as indicated. Results with p_value_ >0.05 are not indicated. Data are presented as median and interquartile ranges. Corrected p_value_ are shown.

### Presymptomatic ALS gene carriers show decreased sleep spindles density

To further characterize sleep microarchitecture defects, we then assessed it in a second prospective cohort study comprising presymptomatic ALS gene carriers, utilising identical inclusion and exclusion criteria. This cohort corresponds to the cohort previously characterized in ^13^, with additional 29 gene carriers and 11 non gene carriers. A total of 57 presymptomatic gene carriers (*SOD1* n=13; *C9orf72* n=33) and 30 first-degree non-carriers relatives (**Table 2**). Similarly to ALS patients, presymptomatic *SOD1* and *C9ORF72* ALS gene carriers demonstrated comparable reductions in sleep spindle density and root mean square (**Figure 2A-B**), with unchanged mean brain-wide amplitude (**Figure 2B**). Density and root mean square of sleep spindles were decreased in both *SOD1* and *C9ORF72* ALS gene carriers in both frontal and motor cortex relevant electrodes (**Figure 2C** ). Interestingly, *SOD1,* but not *C9ORF72,* gene carriers also displayed a mildly decreased amplitude in frontal and motor areas (**Figure 2D-F**). Similar to ALS patients, a brain-wide reduction in slow oscillations and K-complexes was observed in presymptomatic gene carriers (**Supplementary Figures 3 and 4**). Thus sleep microarchitecture is already strongly affected in presymptomatic ALS-gene carriers.

**Figure 2:**
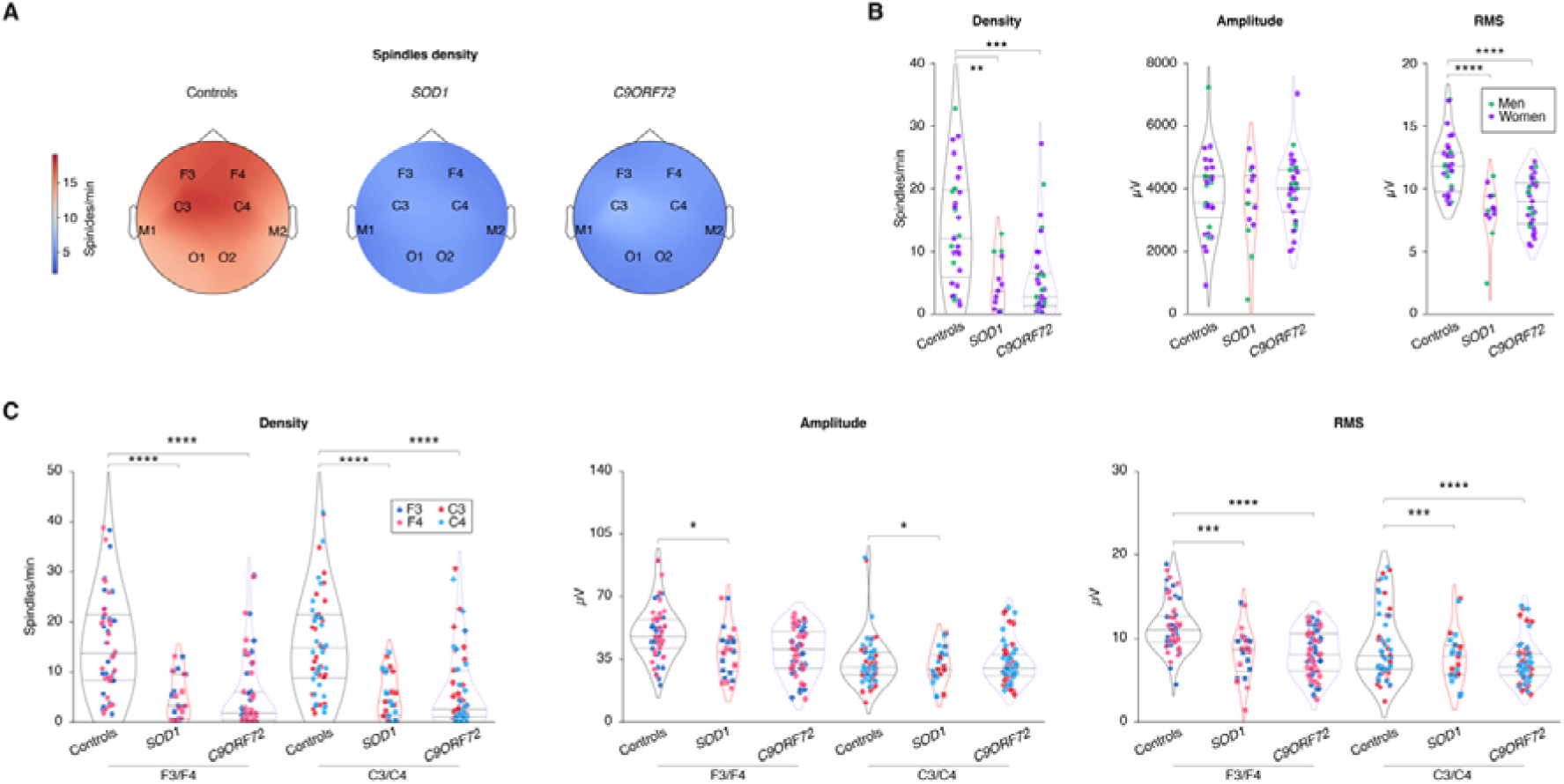
Sleep spindles alterations in presymptomatic ALS gene carriers. **(A)** Topographic map across all electrodes of sleep spindle density in controls, *SOD1* and *C9ORF72* presymptomatic gene carriers. **(B)** Quantification of sleep spindle density, amplitude and root-mean-square (RMS) in controls, *SOD1* and *C9ORF72* presymptomatic gene carriers. Men are shown in green and women in purple. **(C)** Quantification of sleep spindle density, amplitude and root-mean-square (RMS) in controls, *SOD1* and *C9ORF72* presymptomatic gene carriers across F3/F4 and C3/C4 electrodes as indicated. Results with p_value_ >0.05 are not indicated. Data are presented as median and interquartile ranges. Corrected p_value_ are shown.

**Table 2.** Descriptive statistics of the study population of fALS participants. (SEM: standard error of means; BMI: body mass index. Non-parametric Kruskal-Wallis’ test.

|  |  | Gene Carriers | Controls | p<sub>value</sub> | Reference |
| --- | --- | --- | --- | --- | --- |
| Gender | Women (%) | 20 (71.4) | 14 (73.7) |  | 13 |
|  | Men (%) | 8 (28.6) | 5 (26.3) |  |  |
| BMI Mean (±SEM) |  | 26.88 (±0.89) | 26.06 (±1.59) | 0.72 |  |
| Age (±SEM) |  | 40.66 (±2.88) | 41.84 (±2.67) | 0.81 |  |
| Gender | Women (%) | 20 (69) | 7 (63.6) |  | This study |
|  | Men (%) | 9 (31) | 4 (36.4) |  |  |
| BMI Mean (±SEM) |  | 25.62 (±1.30) | 26.94 (±2.172) | 0.629 |  |
| Age (±SEM) |  | 40.71 (±3.04) | 43 (±4.37) | 0.71 |  |

### Microarchitectural alterations of sleep patterns correlate with cognitive deficits

Alterations in sleep microarchitecture are commonly associated with cognitive deficits^14,15^. To determine whether this would be the case in our two cohorts, we correlated sleep microarchitecture parameters with cognitive function in both ALS patients and presymptomatic gene carriers, and with motor function in ALS patients. Cognitive function was evaluated using the Edinburgh Cognitive and Behavioural ALS Screen (ECAS) and motor function with the revised ALS functional rating scale (ALS-FRSr). In ALS patients, a significant positive correlation after adjustment for multiple comparisons (**Figure 3A, Supplementary Table 1**) was observed between sleep spindle density and memory, speech and verbal fluency subscores, as well as the total ECAS score, but not with the ALS-FRS score or its slope (**Figure 3A-E**). Similar correlations, although weaker, were observed in presymptomatic gene carriers between sleep spindle density and memory and executive functions subscores as well as the total ECAS score (**Figure 3F-H, Supplementary Table 2**). Similarly, positive correlations were identified between slow oscillations and total or ECAS subscores, in ALS patients and presymptomatic gene carriers (**Supplementary Figure 5-6**). There were much weaker correlations between cognitive evaluation results and K-complexes in ALS patients but not presymptomatic gene carriers (**Supplementary Figure 6**). Therefore, alterations in the microarchitecture of sleep patterns are associated with cognitive performance, particularly in relation to verbal fluency, speech and memory.

**Figure 3:**
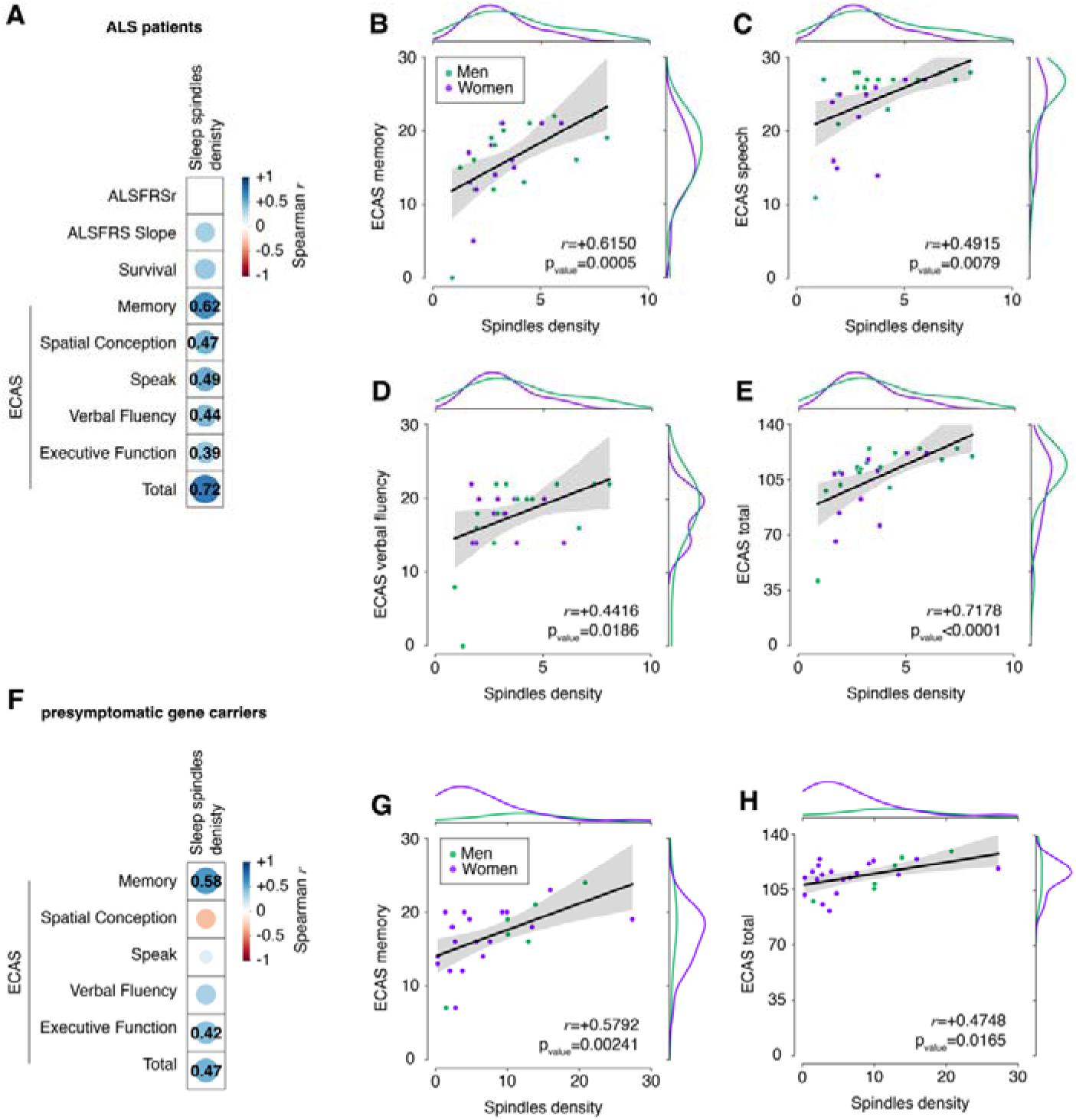
Correlation analysis between sleep microarchitecture and cognitive function in ALS patients. (**A**) Correlation matrix showing Spearman correlation coefficient *r* for each of the corresponding correlations performed in ALS patients. Sleep spindle density was correlated with ALSFRSr, ALSFRS slope, patients’ survival and ECAS subscores as well as the total score for all ALS patients. Only significant correlations are indicated, with the numerical value of the Spearman *r*. (**B-E**) Correlation between sleep spindle density and ECAS memory subscore (**B**) or speech subscore (**C**), verbal fluency subscore (**D)** or total ECAS score (**E**). (**F**) Correlation matrix showing Spearman correlation coefficient *r* for each of the corresponding correlations performed in presymptomatic gene carriers. Sleep spindle density was correlated with ECAS subscores as well as the total score for all *SOD1* and *C9ORF72* gene carriers. Only significant correlations are indicated, with the numerical value of the Spearman *r*. (**G-H**) Correlation between sleep spindle density and ECAS memory subscore (**G**) or total ECAS score (**H**) in presymptomatic gene carriers. In all panels, men are shown in green and women in purple. Spearman pvalue was adjusted with FDR-BKY correction. Spearman correlation coefficient *r* and corrected pvalue are indicated. Side distribution represents sex distribution across both variables (men in green, women in purple).

### Three ALS models exhibit microarchitectural alterations of sleep patterns

Given the presence of sleep microarchitecture alterations in both early ALS patients and presymptomatic gene carriers, we sought to investigate whether these microarchitectural alterations are mirrored by findings in transgenic ALS mouse models. We studied three models expressing different ALS-causing mutations and markedly disparate disease progression. The transgenic model *Sod1^G86R^* is associated with severe and rapidly progressive motor symptoms^16,17^, whereas the *Fus^ΔNLS/+^* ^18,19^ or *TDP-43^Q331K^* models^20^ are linked to a light-to-mild and late-onset phenotype^13^. We used datasets previously acquired^13^ in mouse cohorts implanted with intra-cortical electrodes, and recordings during the presymptomatic phase. As observed in ALS patients and gene carriers, all three models demonstrated substantial alterations in sleep spindles (**Figure 4A-E, Supplementary** Figure 7). It is noteworthy that there was a significant decrease of sleep spindle in 3-month-old *Fus^ΔNLS/+^* mice (**Figure 4C**), while there was no defect in sleep macroarchitecture identified in these animals^13^. As in ALS patients and presymptomatic gene carriers, decreased sleep spindle density was also accompanied by decreased densities in both slow oscillations and K-complexes (**Supplementary Figure 8**). In all three models, microarchitectural alterations were consistently observed in both males and females. Thus early sleep microarchitecture alterations similar to those observed in early ALS patients and gene carriers are observed in multiple ALS mouse models.

**Figure 4:**
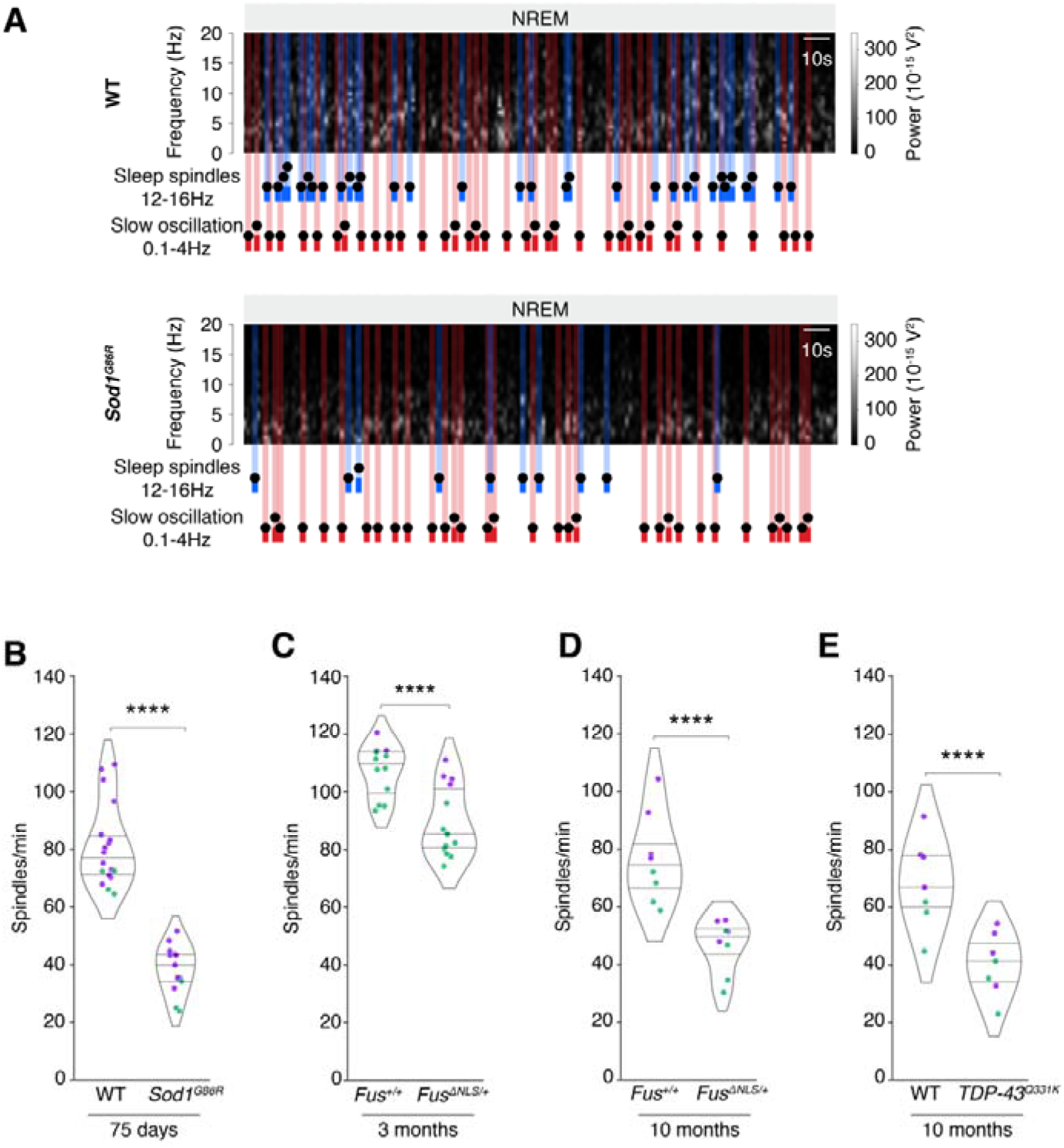
Sleep microarchitecture alterations in *Sod1^G86R^*, *Fus^ΔNLS/+^* and *TDP-43^Q331K^* mice. (**A**) Representative spectrogram of *Sod1^G86R^* mice and their non-transgenic wild-type (WT) littermates at 75 days of age (prior to motor symptom onset). Sleep spindles are labelled in blue and slow oscillation in red on the spectrogram. (**B-E**) Quantification of sleep spindle density in *Sod1^G86R^*mice and their non-transgenic WT littermates at 75 days of age (**B**), in *Fus^ΔNLS/+^* mice and their WT littermates (*Fus^+/+^*) at 3 months of age (**C,** prior to motor symptom onset) or at 10 months of age (**D**) and in TDP-43^Q331K^ mice at 10 months of age (**E**). Independent Student’s t-test with Welch’s t-test with FRD-BKY correction; Data are presented as median and interquartile ranges. Corrected p_value_ are shown.

### Sleep spindle defects are rescued by an orexin antagonist

We previously showed that sleep defects in ALS mouse models can be fully rescued by administration of suvorexant, a dual orexin receptor antagonist. We re-analyzed the previous datasets in which we administered Suvorexant or its vehicle orally at the onset of the inactive period (ie during the day in mice). Acute administration of Suvorexant rescued sleep spindles density in all three mouse models, with a more pronounced effect in females, and decreased efficacy in aged 10-month-old mice (**Figure 5, Supplementary** Figure 9) as well as slow oscillations and K complex loss (**Supplementary Figure 10**). MCH had similar, yet blunted effects on all sleep microarchitecture parameters in either *Sod1*^G86R^ mice (**Supplementary Figure 11**) or *Fus^Δ^*^NLS/+^ mice (**Supplementary Figure 12**). In all, our results suggest that increased orexinergic tone is causally related to sleep microarchitectural defects in both ALS mouse models and patients.

**Figure 5:**
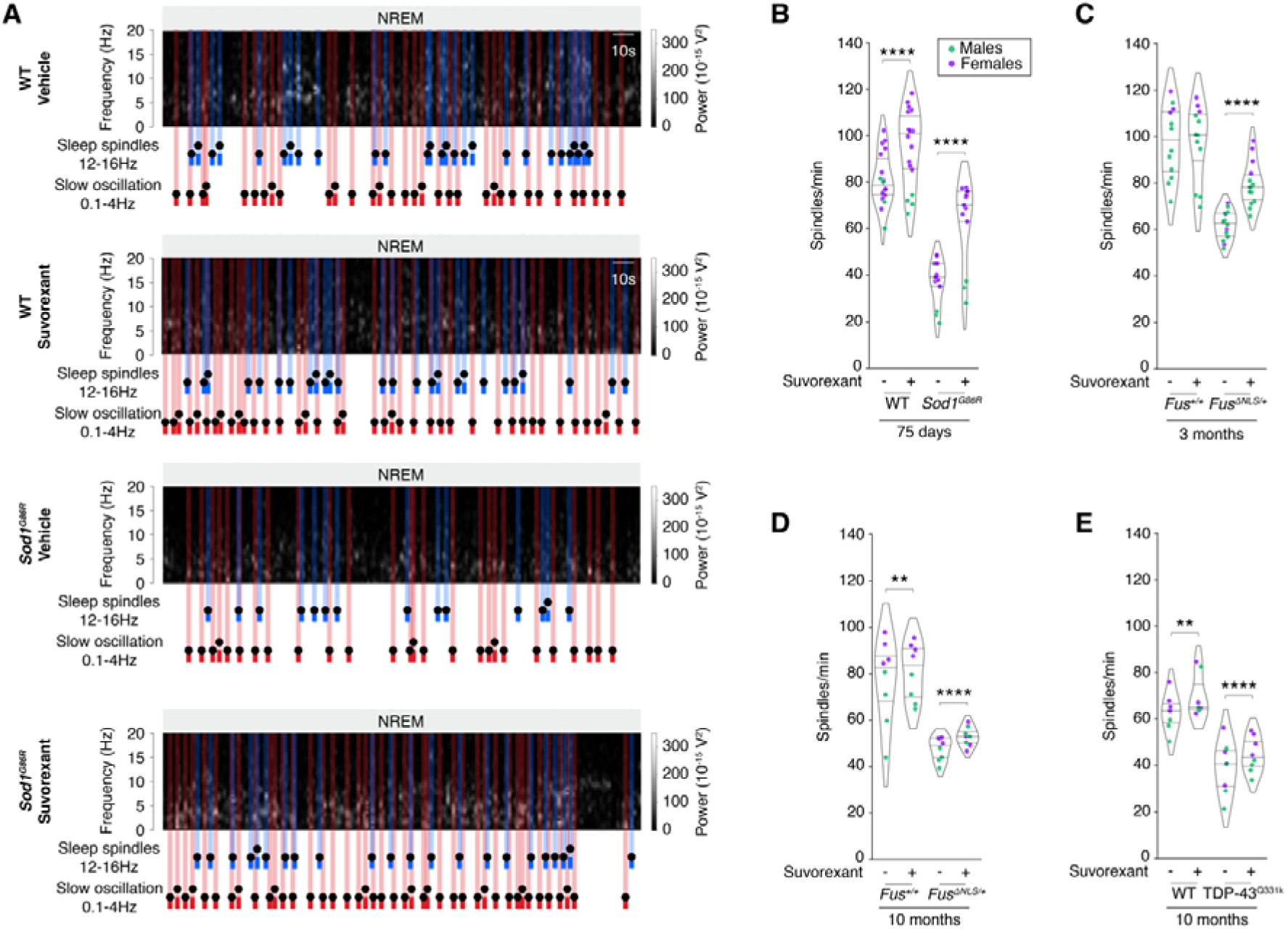
Rescued sleep microarchitecture by Suvorexant *Sod1^G86R^*, *Fus^ΔNLS/+^* and *TDP-43^Q331K^* mice. (A) Representative spectrogram of *Sod1^G86R^*mice and their non-transgenic wild-type (WT) littermates at 75 days of age (prior to motor symptom onset) administered with either vehicle or Suvorexant. Sleep spindles are labelled in blue and slow oscillation in red on the spectrogram. (**B-E**) Quantification of sleep spindle density in mice treated with either vehicle or suvorexant. Genotypes studied include *Sod1^G86R^* mice and their non-transgenic WT littermates at 75 days of age (**B**), in *Fus^ΔNLS/+^*mice and their WT littermates (*Fus^+/+^*) at 3 months of age (**C,** prior to motor symptom onset) or at 10 months of age (**D**) and in TDP-43^Q331K^ mice at 10 months of age (**E**) ns adj. pvalue>0.05, Two-Way ANOVA with Dunn’s test and FDR-BKY correction. Data are presented as median and interquartile ranges. Corrected pvalue are shown.

## Discussion

Here, we show that disruption of sleep microarchitecture is profound, brain-wide and precedes the onset of motor symptoms in ALS, both in humans and in mouse models.

Disruption of sleep microarchitecture involves in ALS involves most sleep-related EEG hallmarks, including sleep spindles, slow oscillations and K-complexes. For all these 3 EEG signals, we observed decreased densities in ALS patients without respiratory impairment, but also in presymptomatic ALS gene carriers, as compared to their respective controls. Decreased density was brain-wide, and we did not observe patterns of disruption related to motor or frontal involvement. We also observed a similar alteration in sleep microarchitecture in 3 different mouse models of ALS. Importantly, defects in sleep microarchitecture were stronger and observed earlier in mouse models than macroarchitectural defects^13^. Indeed, while we previously did not observe increased wake or decreased NREM at 3 months of age in *Fus^ΔNLS/+^* mice, there was already at this age a severe loss of sleep spindles, slow oscillations and K-complexes.

Disruption of sleep EEG hallmarks, as we describe here in ALS, is widely observed in neurological and neurodegenerative diseases. Loss of sleep spindles, especially decreased density, and of slow oscillation, has been repeatedly observed in Alzheimer’s disease ^21–23^ or Parkinson’s disease^24–26^, as well as in other neurological diseases such as temporal lobe epilepsy^27^, schizophrenia^28–30^. This is however not a universal feature of neurological impairment as it is not observed in patients with ADHD, PTSD or most patients with autism spectrum disorder^28^. Interestingly, sleep spindle density also decreases with age^31^, permitting the interpretation that accelerated brain aging could account for some of our results in ALS patients either for microarchitecture defects (the current study) or for macroarchitecture defects^13^. Since decreased spindle density is observed in multiple neurological and neurodegenerative conditions, our current observations are likely not of diagnostic relevance. However, sleep spindles and slow oscillations are quantifiable outcomes, and their defects appear very early in disease. It is thus possible that sleep EEG hallmarks should be investigated for prognostic purposes. Our current lack of information on EEG dynamics during phenoconversion and disease progression hampers evaluation of these alterations as prognostic biomarkers. Future studies should include longitudinal polysomnography in presymptomatic gene carriers to determine the kinetics of sleep macro- and micro-architecture and their possible worsening as a prognostic marker of phenoconversion.

What could be the consequences of defects in sleep microarchitecture? It is widely documented that sleep spindles and slow oscillations are causally involved in memory consolidation and executive functions^14,15^. Indeed, in patients with Parkinson’s or Alzheimer’s diseases, the extent of sleep spindles defects was related to memory and cognitive impairments^21–26^. Complementary to this, we observed strong correlations between cognitive scores and sleep spindles or slow oscillations in our two cohorts. These correlations were more robust than correlations between sleep stages and cognitive function previously observed in the same cohorts^13^. It is currently unknown whether loss of sleep spindles might affect motor progression in ALS. It is however noteworthy that sleep spindles are highly correlated with motor adaptation ^32^ and motor learning ^33–36^, and it is possible that loss of this functional homeostasis might exacerbate motor progression. Longitudinal studies in patients and gene carriers, as well as experimental studies in rodents might address this question.

Sleep spindles originate in the thalamus ^14,15^, and their disruption in ALS indirectly suggests a disruption of cortico-thalamic networks. This disruption could be direct, and caused by intrinsic alteration of thalamic neurons or thalamo-cortical pathways. Atrophy of the thalamus has been largely documented in ALS, in particular in *C9ORF72* patients ^37–42^, and alterations of thalamo-cortical pathways has been observed in sporadic ALS ^43,44^ and in presymptomatic *C9ORF72* gene carriers ^45^. These thalamic alterations have been related to faster progression in sporadic ALS ^46^ and higher risk of phenoconversion in *C9ORF72* ALS ^47^. Thalamic involvement has been considered an early biomarker of ALS, that contributes to motor and cognitive deficits in sALS ^48–51^. It is also possible that these defects are indirectly caused by abnormal signaling by hypothalamic neuropeptides, such as orexin. Indeed, thalamic reticular nucleus neurons are sensitive to orexin ^52,53^, and we provide evidence that orexin antagonism or MCH supplementation are able to rescue the loss of sleep spindles and slow oscillation in several ALS mice. This is consistent with the loss of MCH neurons observed in ALS^54^, and several studies pointing to orexin alterations in ALS patients and mouse models^13,55,56^. It is also possible that defects in other neuromodulators, in particular acetylcholine^57–60^, cause these defects in sleep microarchitecture, which would be consistent with previous observations on acetylcholine defects in ALS^60^. Defects in norepinephrine could also cause these defects in sleep microarchitecture as this neurotransmitter is highly involved in sleep-related cognitive consolidation^61–63^. This would be consistent with our previous observations on norepinephrine defects in ALS^64^. Whether defects in sleep microarchitecture directly originate from degeneration or dysfunction of thalamic reticular nucleus neurons or from indirect circuit dysfunction involving direct LHA innervation or other neuronal relays will require further experimental work.

Summarizing, our current work establishes disruption of sleep EEG as a very early biomarker of ALS, detectable many years before onset of symptoms in presymptomatic gene carriers and correlated with cognitive impairment. Elucidation of the underlying mechanisms might shed light on the very precocious events in ALS pathophysiology, and the relevance of these alterations for prognosis should be studied longitudinally in prospective cohorts.

## Materials and Methods

### Patients/participants

ALS patients were recruited from the inpatient and outpatient clinics of the neurologic department of the University Hospital of Ulm, Germany. The inclusion criteria for ALS patients included a diagnosis of definite ALS based on the revised El Escorial criteria[30]. Presymptomatic carriers of fALS genes were recruited through the study centre of the Neurological University Hospital, through which first-degree relatives of confirmed familial ALS patients receive longitudinal follow-up and counselling. Controls were recruited from the general population at the neurology clinic, and matched to ALS patients based on age, sex, and geographical location; the requirement for this group was the exclusion of neurodegenerative diseases. All individuals in the control group were unrelated to ALS or familial ALS.

The study in the ALS patient cohort was approved by the Ethics Committee of the University of Ulm (reference 391/18), as well as the study in the presymptomatic carriers which was also approved by the Ethics Committee of the University of Ulm (reference 68/19), in compliance with the ethical standards of the current version of the revised Helsinki Declaration. All participants gave informed consent prior to enrolment.

Medical history was documented. For ALS patients, the ALSFRS-r and characteristics of disease progression were documented (site of first paresis/atrophy, date of onset). All participants also completed validated daytime sleepiness and sleep quality questionnaires, namely the Epworth Sleepiness Scale (ESS) ^65^ and the Pittsburgh Sleep Quality Index (PSQI) ^66^.

### Patients’ inclusion process

The same exclusion criteria employed by Guillot SJ *et al.,*^13^ were used. The exclusion criteria were intended to exclude all possible circumstances that might otherwise alter sleep architecture. For this reason, participants who had an apnoea-hypopnea index (AHI) above 20 per hour or participants who had a periodic limb movement index (PLMSI) above 50 per hour were excluded. In particular, we intended to exclude respiratory insufficiency in ALS patients. Respiratory insufficiency develops earlier or later in the progression of ALS, depending on the individual course, but is generally present in advanced stages, and is known to influence sleep architecture. For this reason, ALS patients received transcutaneous capnometry in addition to polysomnography.

### Neuropsychological Assessment

Cognition was measured with the German version of the Edinburgh Cognitive and Behavioural ALS Screen (ECAS) ^67–69^ by trained neuropsychologists. The ECAS addresses cognitive domains of language, verbal fluency, executive functions (ALS-specific functions) and memory and visuospatial functions (ALS non-specific functions). Age and education-adjusted cut-offs were used^69^. Behavioural changes were assessed by patient caregiver/1st-degree relative interviews on disinhibition, apathy, loss of sympathy/empathy, perseverative/stereotyped behaviour, hyperorality/altered eating behaviour and psychotic symptoms.

### Electroencephalography in patients and subjects

All participants, ALS patients, healthy controls, fALS gene carriers and fALS controls underwent a 1-night full Polysomnography, involving monitoring of various physiological parameters including electroencephalogram (EEG), surface electromyogram (EMG), electrooculogram (EOG) respiratory effort and flow, pulse and oxygen saturation. All measurements were conducted according to the criteria of the American Academy of Sleep Medicine (AASM) guidelines^70,71^. The EEG electrodes were placed according to the international 10-20 system, the following electrodes were used in each subject: Fz, C3, C4, Cz, P3, P4, Pz, O1, O2, A1, and A2. The sampling rate was 512 Hz in each case. The individually different point in time at which the participant turned off the lights and tried to sleep was marked with a “lights off” marker in each recording.

### Sleep analyses in patients and subjects

Analyses were performed using available Python packages (only compatible with Python 3.10 or newer, Python Software Foundation. Python Language Reference, version 3.12. Available at http://www.python.org) relying on MNE package[38]. EEGs preprocessing was performed following section *res2hsleep*.

Briefly, recordings were first de-identified using the open-source Prerau Lab EDF De-identification Tool (Version 1.0; 2023) in Python (Prerau Lab EDF De-identification Tool [Computer software], 2023, Retrieved from https://sleepeeg.org/edf-de-identification-tool), to then be notch-filtered to remove the 50Hz powerline. Independent component analysis was performed to remove all remaining artefacts from the signal^72–76^.

Analyses of sleep spindles, slow oscillations and K-complexes were performed on all electrodes^77–79^ and outliers were removed using an isolation forest algorithm^80^. K-complexes analysis was limited to the sensorimotor cortex (C3), which is known to be impaired in ALS, using MNE and SciPy packages^80,81^. For the sleep spindle, its density (number of sleep spindles per minute of NREM 2 sleep), amplitude (peak-to-peak amplitude of the detrended sleep spindle) and root-mean-square (RMS) were analysed. For the slow oscillation, its density (number of slow oscillations per minute of NREM2), slope (slope between the negative peak and the mid-crossing of the slow oscillation), phase-amplitude coupling (PAC; slow oscillations-sleep spindles normalised PAC within a 2sec epoch centred around the negative peak of the slow oscillation) and phase at Sigma peak (slow oscillation’s phase when sigma peak is reached within a 2sec epoch centred around the negative peak of the slow oscillation) were analysed. For the K-complex assessment, we analysed its density (number of K-complexes per minute of NREM 2).Topographic maps were performed using MNE and YASA packages ^82^. All analyses were performed following the AASM’s guidelines^83^.

### Mouse models

All experiments were performed in strict compliance with Directive 2010/63/EU, and new Regulation (EU) 2019/1010, and the project was reviewed and approved by the Ethics Committee of the University of Strasbourg and the French Ministry of Higher Education, Research and Innovation (Decree n°2013-118, February 1^st^, 2013). All datasets, animal care, surgery and procedures used in this study have been previously described in ^13^

### Electrocorticography analysis in mice

Data were extracted from NeuroScore^™^ software for sleep and seizure analysis 3.4 (Data Science International Inc., St. Paul, MN, USA) and used in combination with already available Python packages (Python Software Foundation. Python Language Reference, version 3.12. Available at http://www.python.org) to further process the data.

Sleep spindles, slow oscillations and K-complexes were automatically detected based using publicly available pipelines ^84,85^.The signal was first band-pass filtered at 1-45Hz, and the sigma power (12-16Hz) was calculated on a 200ms Hamming window followed by a Short-Term Fourier Transform (STFT) with the same window length. The occurrence of sleep spindles was identified when the smoothed absolute sigma power within the 12-16 Hz range exceeded 0.2 of the total power observed in the broadband frequency range of 0.1-45Hz. This signifies that a minimum of 20% of the signal’s total power must be contained within the specified sigma band.

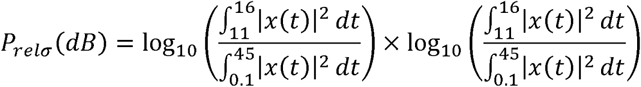

For slow oscillations, the signal was first band-pass filtered at 0.1-45Hz, and the low delta power (0.1-2Hz) was calculated on a 400ms Hamming window. Areas under the curve (AUC) were calculated using Simpson’s rule derived from the delta band (A_SO_) and the total power broadband frequency range (B_PSD_ ). The ratio of these two AUCs was then obtained, providing the slow oscillations ratio.

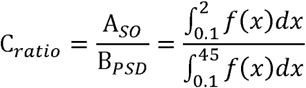

For K-complexes, the signal was first band-pass filtered at 0.1-45Hz, and the sigma power was calculated on a 400ms Hamming window. The signal was first band-pass filtered at 1-45Hz, and the low delta power (0.3-1Hz) was calculated on a 200ms Hamming window followed by an STFT with the same window length. The occurrence of K-complexes was identified when the STFT within the 0.3-1Hz range exceeded the mean STFT of the same frequency range.

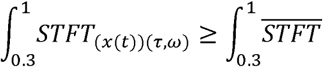

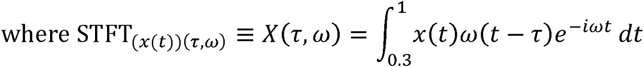

### Statistical analyses

G*Power software (Version 3.1.9.6 for macOS; 2023) was used to determine the sufficient sample size needed to reach significant statistical power using an *A priori* Student’s t-test coupled with a linear bivariate regression ^86,87^.

Prior to any statistical analysis, normality and homoscedasticity were both tested respectively with Shapiro-Wilk test ^88^ and Bartlett’s test^89^.

Statistical analysis of two groups was performed using an independent Student’s t-test, from Pingouin ^90^, using the Welch t-test correction, from SciPy, as recommended by Zimmerman ^91^ and with a large Cauchy scale factor due to the considerate effect size ^92^. When data were heteroscedastic and normality was not met, a Mann-Withney *U* test was performed using SciPy ^80^.

Follow-up analysis were performed using paired t-test from SciPy^80^ or a Wilcoxon-Mann-Whitney rank-sum test from statsmodels ^93^ when normality was not met. P_values_ were then adjusted using FDR-BKY correction.

For statistical analysis of three or four groups, a One-way ANOVA or Two-way ANOVA was performed using Pingouin ^90^ toolbox. For both One-way ANOVA and Two-way ANOVA, a one-step Bonferroni correction was applied. When data were heteroscedastic and normality was not met, a Kruskal-Wallis from SciPy ^80^ followed by Dunn’s multiple comparison test with FDR-BKY correction was performed using scikit-posthocs ^94^, instead of a One-Way ANOVA. For the Two-Way ANOVA, a generalized least squares model was fitted using statsmodels ^93^, followed by Dunn’s multiple comparison and FDR-BKY correction using scikit-posthocs ^94^. We evaluated whether a sex-specific effect was present in all our analyses by performing a Two-way ANOVA with a one-step Bonferroni correction for both sexes. Sex was self-reported in both ALS cohorts.

Spearman’s correlation coefficient from SciPy^80^, were used to determine correlations on non-parametric data.

Data are presented as violin plots with all points and expressed as average ± interquartile. Plots were generated using Seaborn and Matplotlib packages^95^. Results were deemed significant when their adj. p_value_<0.05. Here, only corrected p_values_ (adj. p_value_) are shown.

## Data Availability

All data produced in the present study are available upon reasonable request to the authors

---

**Supplementary Figure 1:**
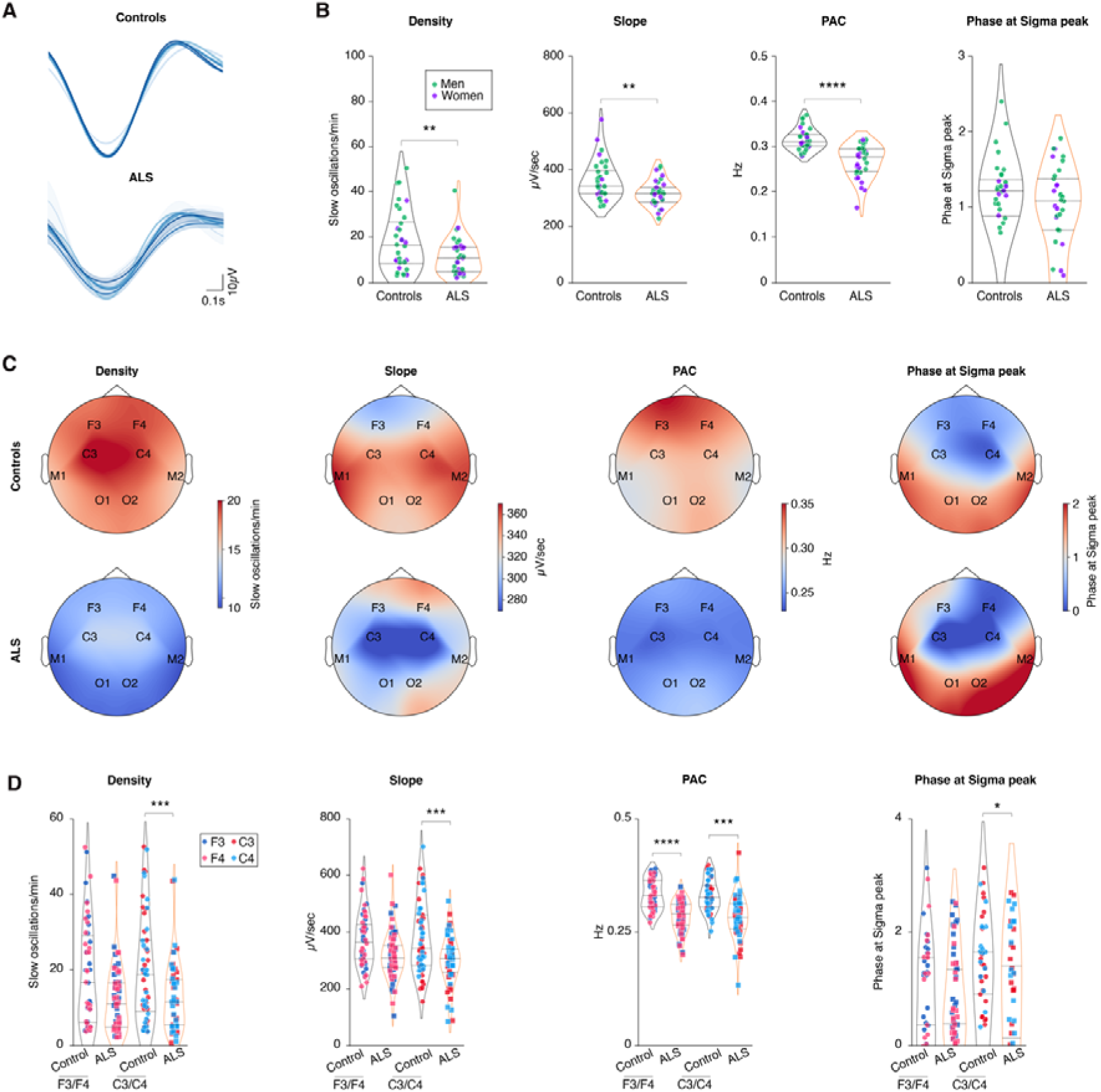
Slow oscillations alterations in early ALS patients. (A) Representative slow oscillations across all electrodes of a healthy individual and one sporadic ALS patient. (B) Quantification of slow oscillation density, slope, phase-amplitude coupling between sleep spindles and slow oscillations (PAC) and phase at Sigma peak (PSP) in controls and ALS patients. Men are shown in green and women in purple. (C) Topographic maps across all electrodes of slow oscillation density, slope, phase-amplitude coupling and phase at Sigma peak in Controls (upper row) and ALS patients (lower row). (D) Quantification of slow oscillation density, slope, phase-amplitude coupling and phase at Sigma peak across F3/F4 and C3/C4 electrodes as indicated. Only p values <0.05 are shown. Data are presented as median and interquartile ranges. Corrected p_value_ are shown.

**Supplementary Figure 2:**
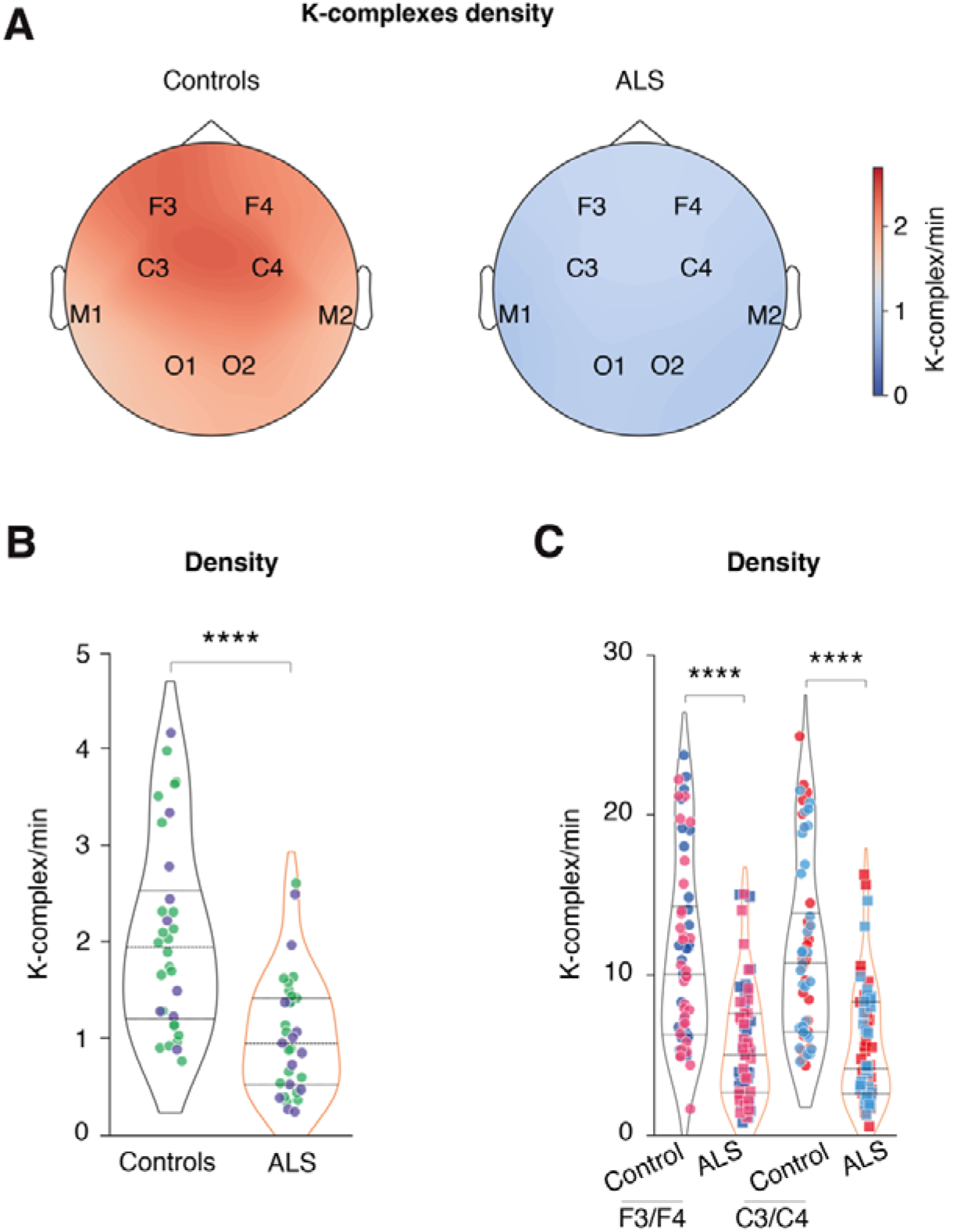
K-complexes alterations in early ALS patients. **(A)** Topographic map across all electrodes of K-complex density in controls and ALS patients. **(B)** Quantification of K-complex density in controls and ALS patients. Men are shown in green and women in purple. **(C)** Quantification of K- complex density across F3/F4 and C3/C4 electrodes as indicated.

**Supplementary Figure 3:**
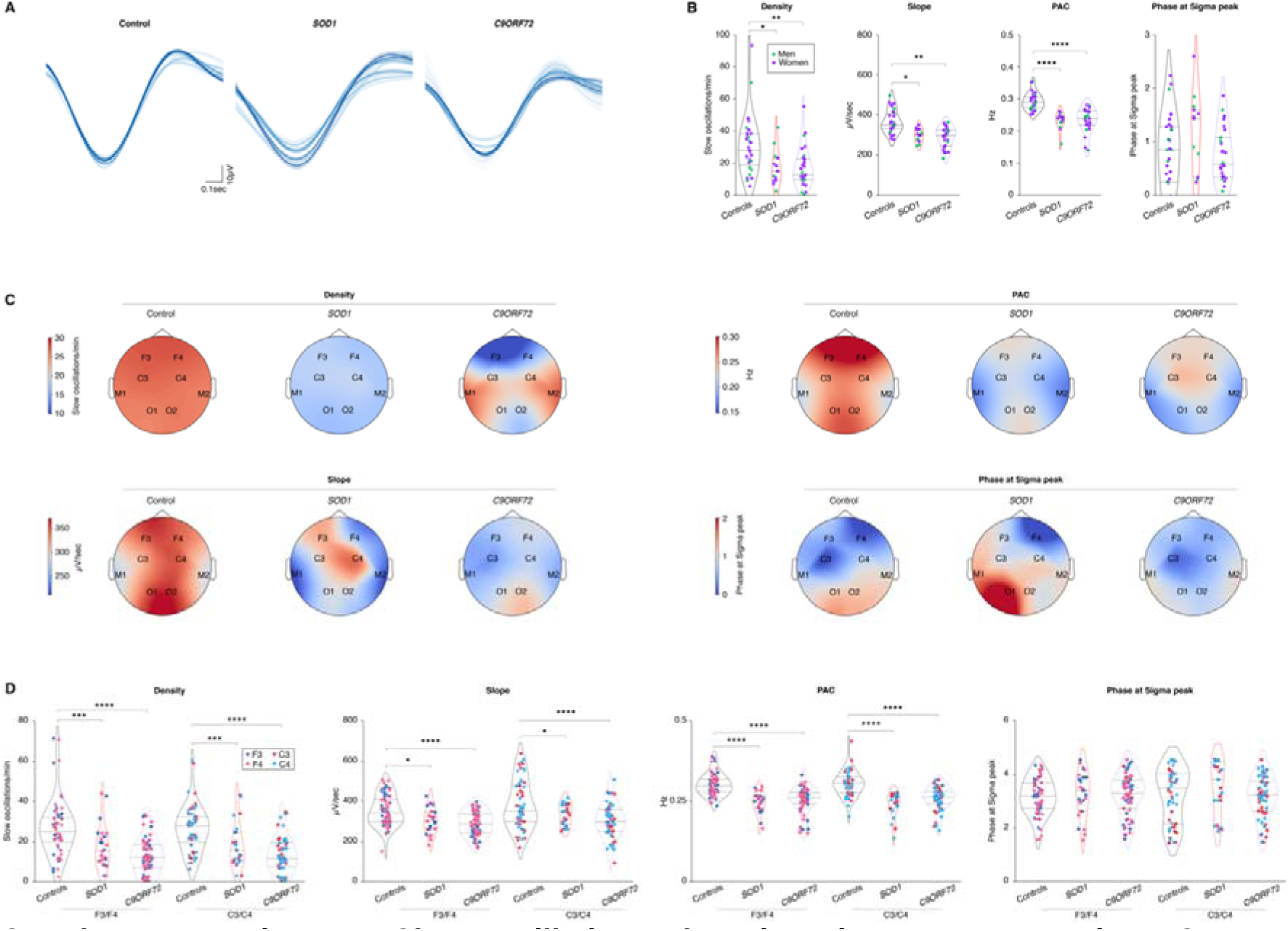
Slow oscillations alterations in presymptomatic ALS gene carriers. (A) Representative slow oscillations across all electrodes of a healthy individual and one presymptomatic *SOD1* and *C9ORF72* gene carrier. (B) Quantification of slow oscillation density, slope, phase-amplitude coupling between sleep spindles and slow oscillations (PAC) and phase at Sigma peak (PSP) in controls, *SOD1* and *C9ORF72* presymptomatic gene carriers. Men are shown in green and women in purple. (C) Topographic maps across all electrodes of slow oscillation density, slope, phase-amplitude coupling and phase at Sigma peak in controls, *SOD1* and *C9ORF72* presymptomatic gene carriers. (D) Quantification of slow oscillation density, slope, phase-amplitude coupling and phase at Sigma peak across F3/F4 and C3/C4 electrodes in controls, *SOD1* and *C9ORF72* presymptomatic gene carriers as indicated.

**Supplementary Figure 4:**
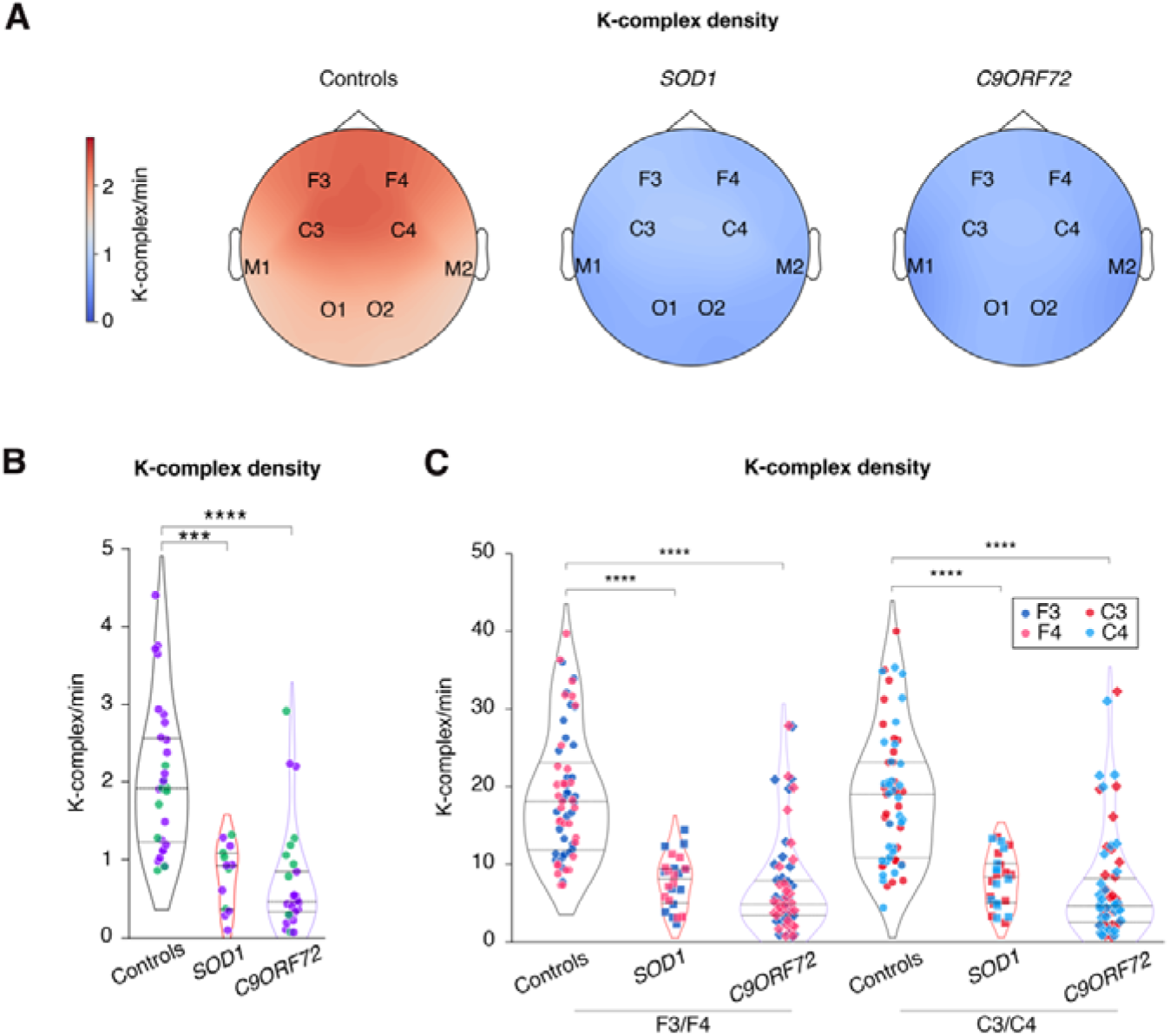
K-complexes alterations in presymptomatic ALS gene carriers. **(A)** Topographic map across all electrodes of K-complex density in controls, *SOD1* and *C9ORF72* presymptomatic gene carriers. **(B)** Quantification of K-complex density in controls, *SOD1* and *C9ORF72* presymptomatic gene carriers. Men are shown in green and women in purple. **(C)** Quantification of K- complex density across F3/F4 and C3/C4 electrodes in controls, *SOD1* and *C9ORF72* presymptomatic gene carriers as indicated.

**Supplementary Figure 5:**
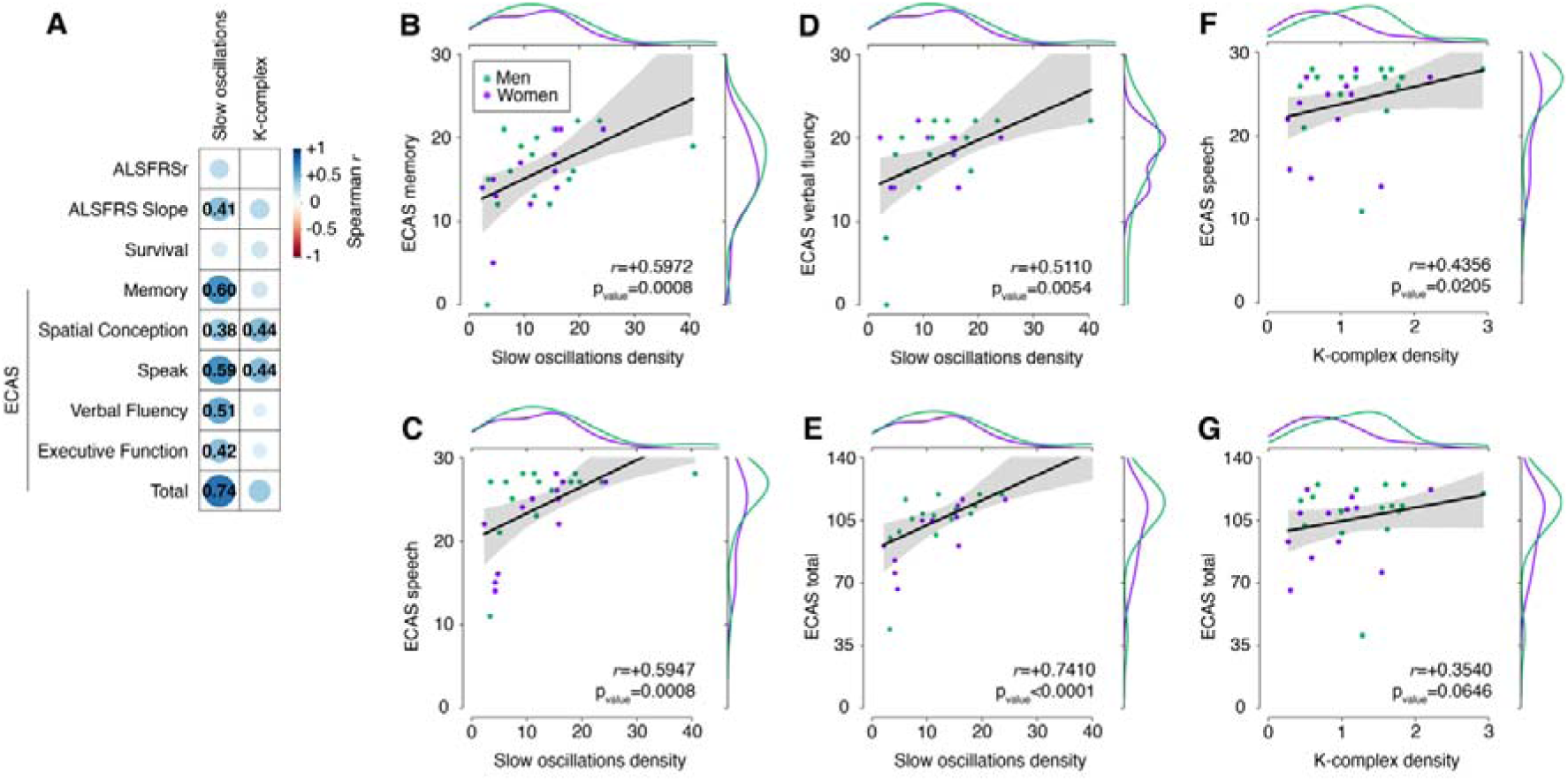
Correlation analysis between slow oscillations and K- complex density and cognitive function in ALS patients. (**A**) Correlation matrix showing Spearman correlation coefficient *r* for each of the corresponding correlations performed in ALS patients. Slow oscillation or K-complex density were correlated with ALSFRSr, ALSFRS slope, patients’ survival and ECAS subscores as well as the total score for all ALS patients. Only significant correlations are indicated, with the numerical value of the Spearman *r*. (**B-E**) Correlation between slow oscillation density and ECAS memory subscore (**B**) or verbal fluency subscore (**C**), speech subscore (**D)** or total ECAS score (**E**). (**F-G**) Correlation between K-complex density and ECAS speech subscore (**F)** or total ECAS score (**G**). In all panels, men are shown in green and women in purple. Spearman pvalue was adjusted with FDR-BKY correction. Spearman correlation coefficient *r* and corrected pvalue are indicated. Side distribution represents sex distribution across both variables (men in green, women in purple).

**Supplementary Figure 6:**
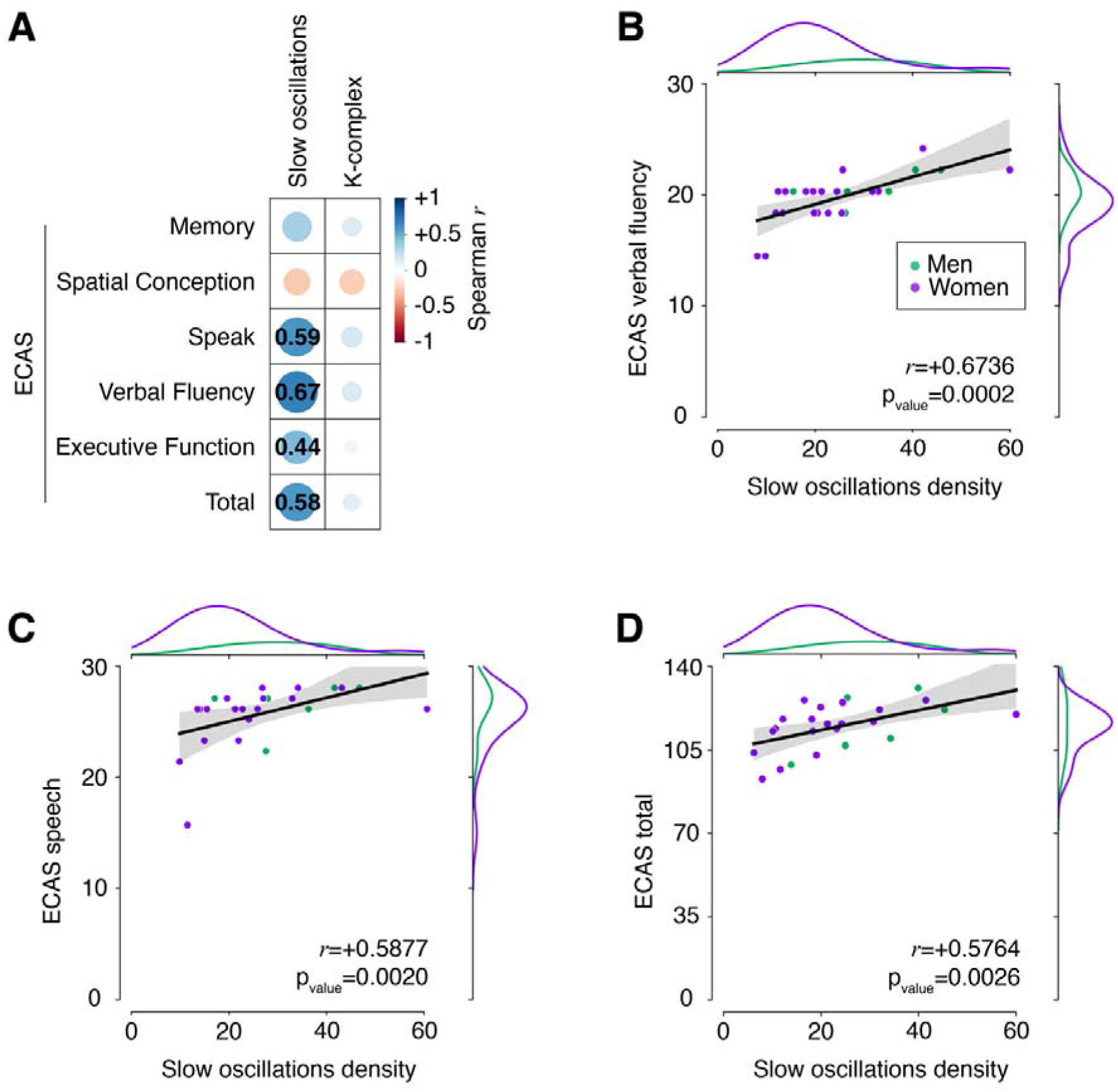
Correlation analysis between slow oscillations and K- complex density and cognitive function in presymptomatic ALS gene carriers. (**A**) Correlation matrix showing Spearman correlation coefficient *r* for each of the corresponding correlations performed in presymptomatic gene carriers. Slow oscillation and K-complex densities were correlated with ECAS subscores as well as the total score for all *SOD1* and *C9ORF72* gene carriers. Only significant correlations are indicated, with the numerical value of the Spearman *r*. (**B-D**) Correlation between slow oscillation density and ECAS verbal fluency subscore (**B**), speech subscore (**C**) or total ECAS score (**D**) in presymptomatic gene carriers. In all panels, men are shown in green and women in purple. Spearman pvalue was adjusted with FDR-BKY correction. Spearman correlation coefficient *r* and corrected pvalue are indicated. Side distribution represents sex distribution across both variables (men in green, women in purple).

**Supplementary Figure 7:**
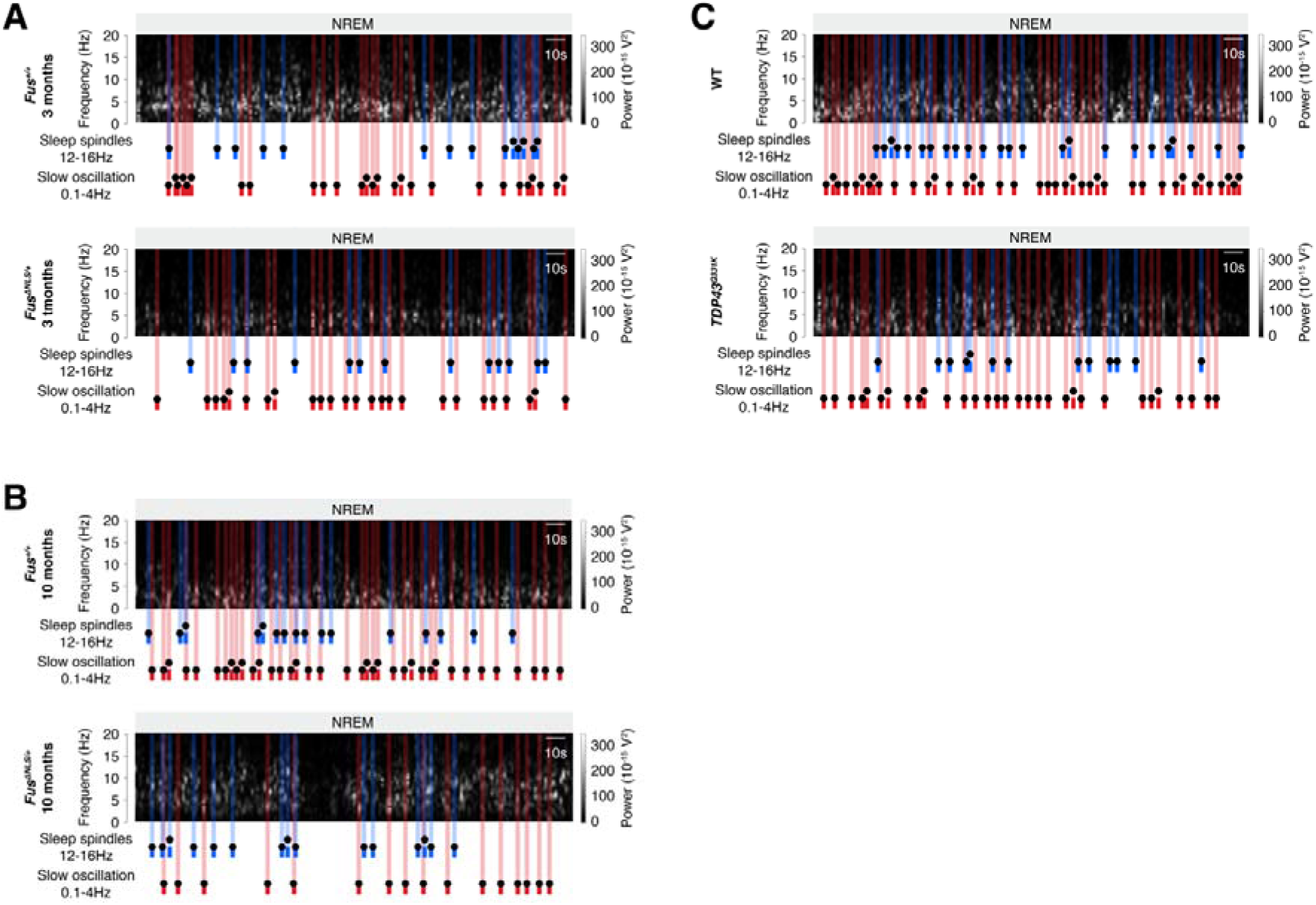
Sleep microarchitecture alterations in *Fus*^Δ*NLS/+*^ and *TDP-43^Q331K^* mice. (**A-C**) Representative spectrogram of mice *Fus^ΔNLS/+^* mice and their WT littermates (*Fus^+/+^*) at 3 months of age (**A,** prior to motor symptom onset) or at 10 months of age (**B**) and in TDP-43^Q331K^ mice at 10 months of age (**C**). Sleep spindles are labelled in blue and slow oscillation in red on the spectrogram.

**Supplementary Figure 8:**
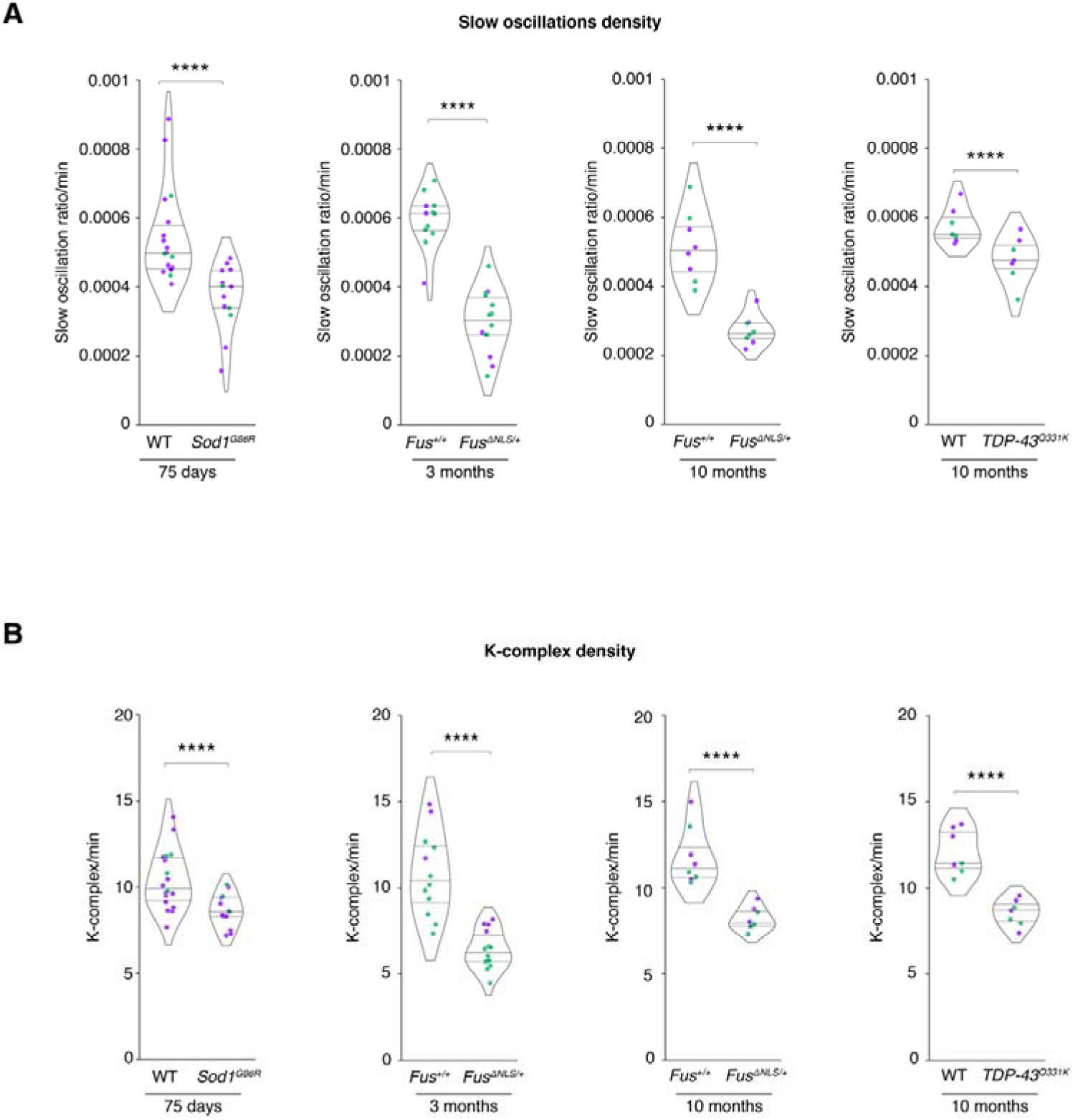
additional sleep microarchitecture alterations in *Sod1^G86R^*, *Fus^ΔNLS/+^* and *TDP-43^Q331K^* mice. (**A-B**) Quantification of slow oscillation density (**A**) and K-complex density (**B**) in *Sod1^G86R^* mice and their non-transgenic WT littermates at 75 days of age, in *Fus^ΔNLS/+^* mice and their WT littermates (*Fus^+/+^*) at 3 months of age (prior to motor symptom onset) or at 10 months of age and in TDP-43^Q331K^ mice at 10 months of age. Independent Student’s t-test with Welch’s t-test with FRD-BKY correction. Data are presented as median and interquartile ranges. Corrected p_value_ are shown.

**Supplementary Figure 9:**
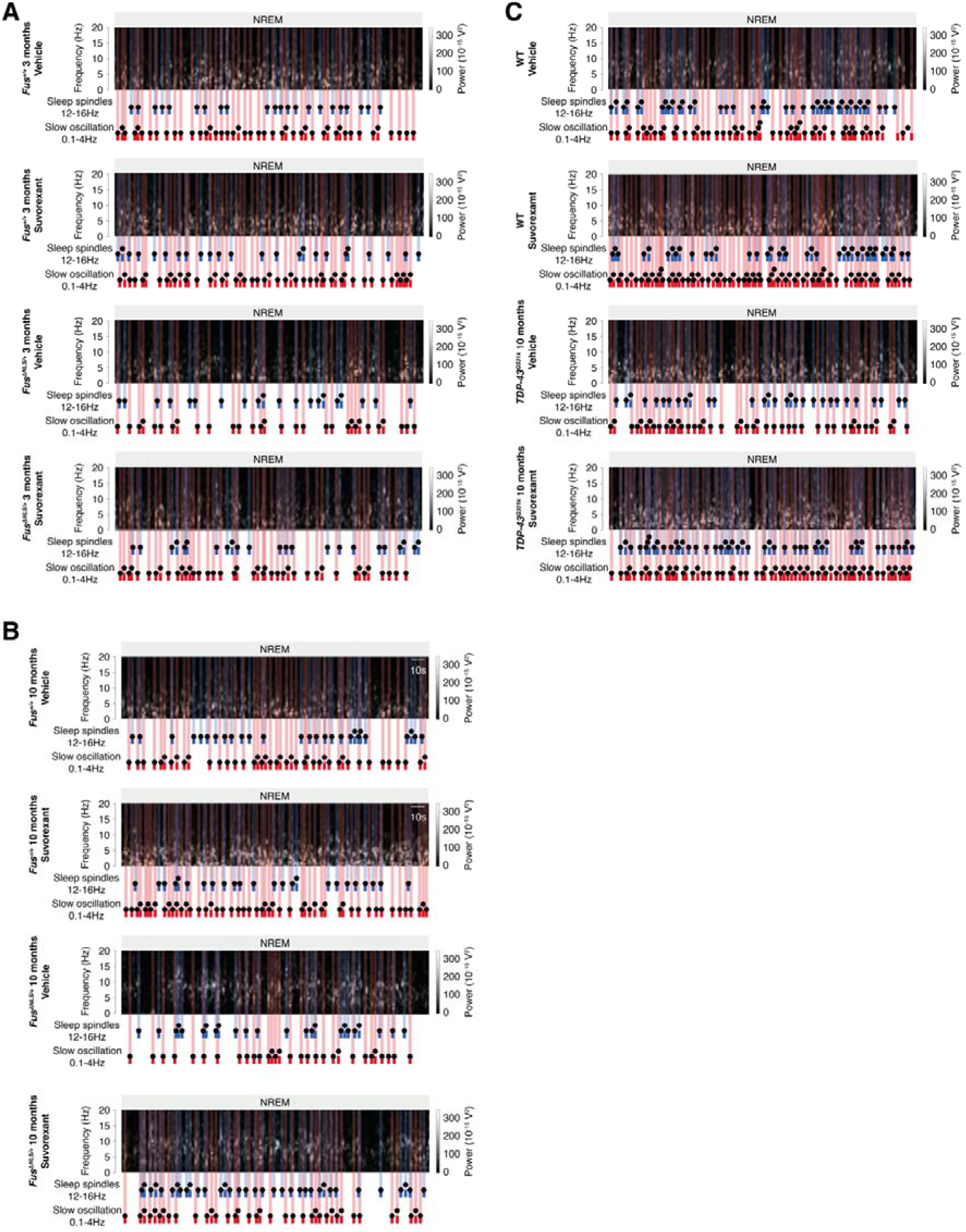
Sleep microarchitecture alterations in *Fus^ΔNLS/+^* and *TDP-43^Q331K^* mice treated with suvorexant. (**A-C**) Representative spectrogram of mice administered with either vehicle or Suvorexant. Representative spectrograms for indicated genotypes are shown: *Fus^ΔNLS/+^* mice and their WT littermates (*Fus^+/+^*) at 3 months of age (**A,** prior to motor symptom onset) or at 10 months of age (**B**) and in TDP-43^Q331K^ mice at 10 months of age (**C**). Sleep spindles are labelled in blue and slow oscillation in red on the spectrogram.

**Supplementary Figure 10:**
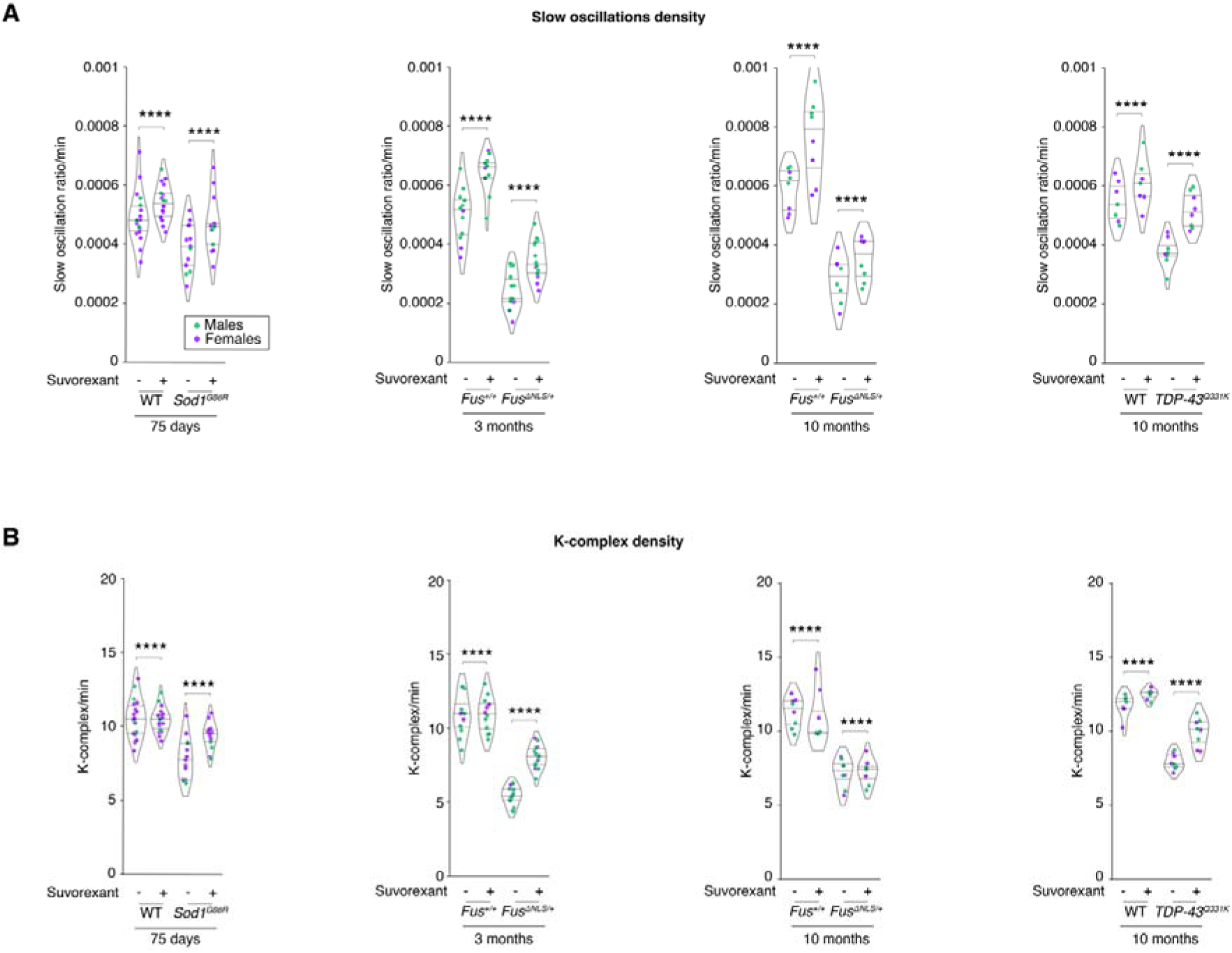
additional results on sleep microarchitecture in *Sod1^G86R^*, *Fus^ΔNLS/+^* and *TDP-43^Q331K^* mice treated with suvorexant. (**A-B**) Quantification of slow oscillation density (**A**) and K-complex density (**B**) in *Sod1^G86R^* mice and their non-transgenic WT littermates at 75 days of age, in *Fus^ΔNLS/+^*mice and their WT littermates (*Fus^+/+^*) at 3 months of age (prior to motor symptom onset) or at 10 months of age and in TDP-43^Q331K^ mice at 10 months of age . Mice were either administered vehicle or suvorexant as indicated. Data are presented as median and interquartile ranges. Corrected p_value_ are shown.

**Supplementary Figure 11:**
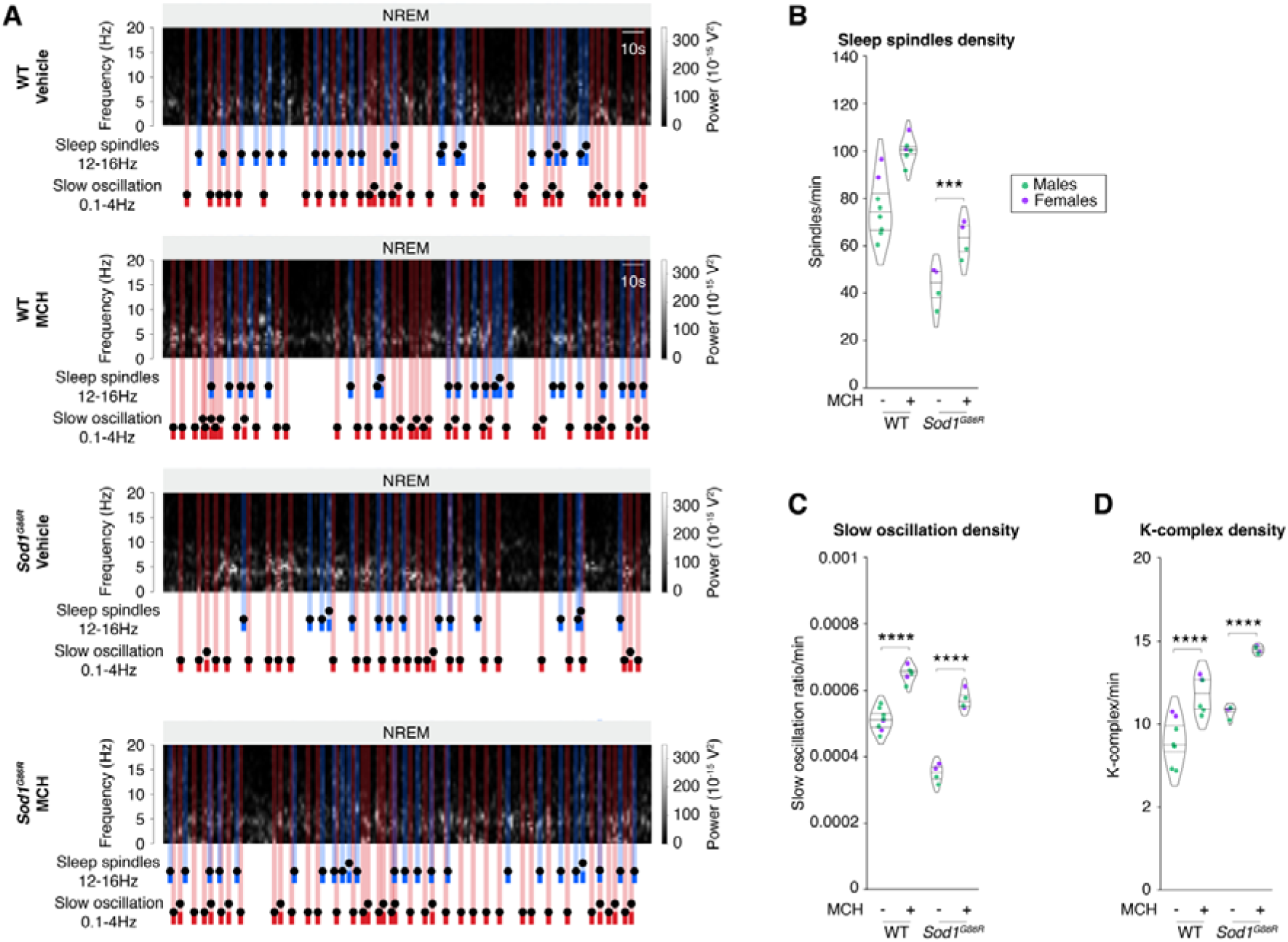
Rescued sleep microarchitecture by MCH chronic delivery in *Sod1^G86R^*mice. (**A**) Representative spectrogram of *Sod1^G86R^* mice and their non-transgenic wild-type (WT) littermates at 75 days of age (prior to motor symptom onset) administered with either vehicle or MCH via icv cannulation and osmotic minipump delivery. Sleep spindles are labelled in blue and slow oscillation in red on the spectrogram. (**B-D**) Quantification of sleep spindle density (**B**), slow oscillation density (**C**) and K-complex density (**D**) in *Sod1^G86R^* mice and their non-transgenic WT littermates treated with either vehicle or MCH at 75 days of age Two-Way ANOVA with Dunn’s test and FDR-BKY correction. Data are presented as median and interquartile ranges. Corrected pvalue are shown.

**Supplementary Figure 12:**
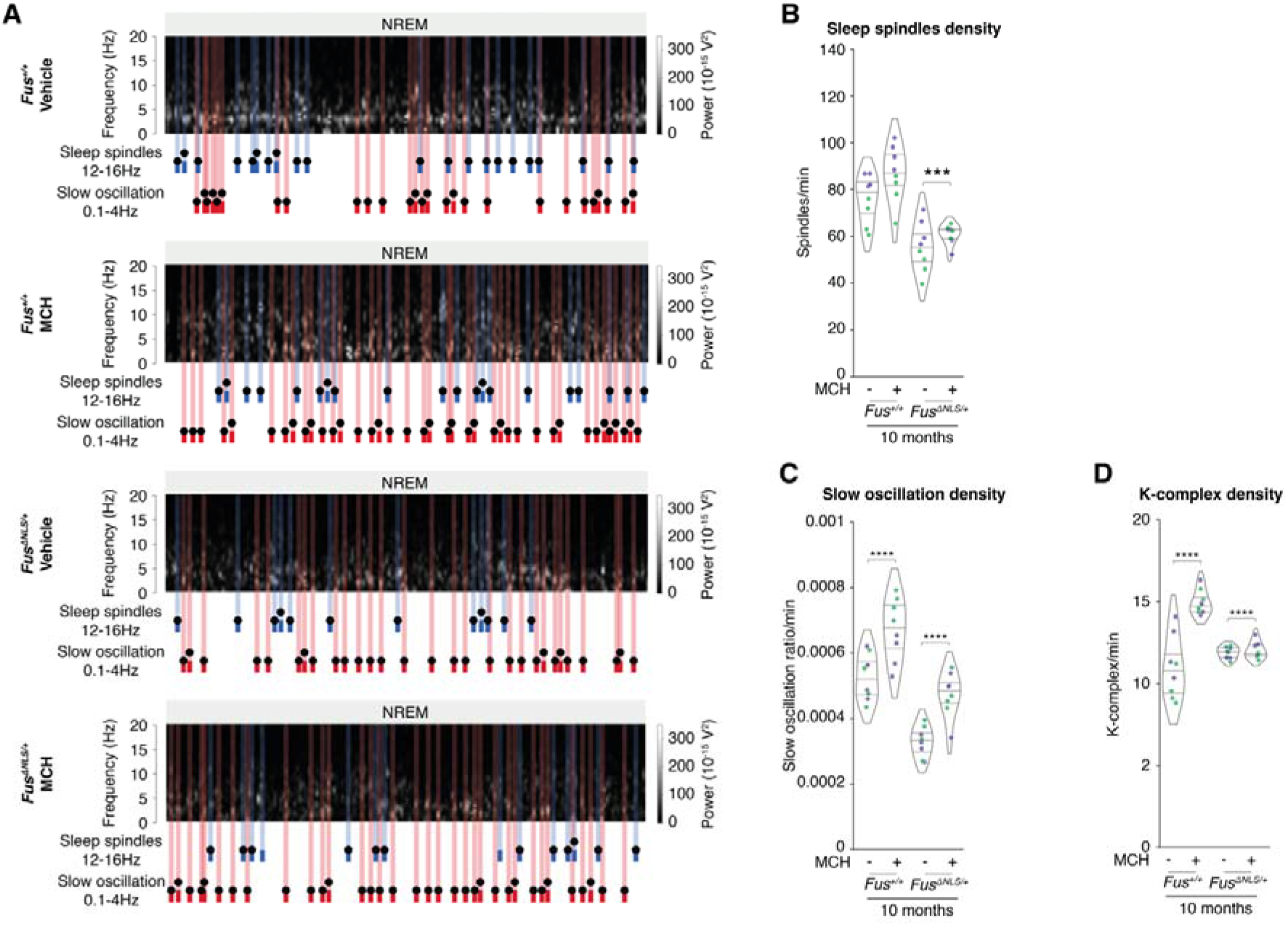
Rescued sleep microarchitecture by MCH chronic delivery in *Fus^ΔNLS/+^* mice. (**A**) Representative spectrogram of *Fus*^Δ*NLS/+*^ mice and their non-transgenic wild-type (WT) littermates at 75 days of age (prior to motor symptom onset) administered with either vehicle or MCH via icv cannulation and osmotic minipump delivery. Sleep spindles are labelled in blue and slow oscillation in red on the spectrogram. (**B-D**) Quantification of sleep spindle density (**B**), slow oscillation density (**C**) and K-complex density (**D**) in *Fus^ΔNLS/+^* mice and their WT littermates (*Fus^+/+^*) mice treated with either vehicle or MCH at 10 months of age. Two-Way ANOVA with Dunn’s test and FDR-BKY correctionData are presented as median and interquartile ranges. Corrected pvalue are shown.

**Supplementary Table 1:** correlation between sleep microarchitecture and clinical parameters in ALS patients. Spearman r and adjusted p-values are indicated for each comparison.

|  | Density of sleep spindles |  | Density of slow oscillations |  | Density of K-complexes |  |
| --- | --- | --- | --- | --- | --- | --- |
| | $r$ | pvalue | $r$ | pvalue | $r$ | pvalue |
| ALSFRS<sub>r</sub> | -0,076277984 | 0,67309535 | 0,3262816 | 0,06386008 | 0,0700615 | 0,69843741 |
| ALSFRS_Slope | 0,333584024 | 0,05780799 | 0,41129774 | 0,01740962 | 0,27893374 | 0,11596195 |
| Survival | 0,374781661 | 0,23000298 | 0,17162898 | 0,59378681 | 0,20315268 | 0,52656305 |
| ECAS_MEM | 0,615054037 | 0,00049579 | 0,59717456 | 0,0007936 | 0,19529891 | 0,31927845 |
| ECAS_SpatConpt | 0,465622028 | 0,01252376 | 0,37872778 | 0,0468773 | 0,43857004 | 0,01956586 |
| ECAS_Speak | 0,491513019 | 0,00790041 | 0,59465291 | 0,00084618 | 0,43563399 | 0,02049491 |
| ECAS_VerFlnt | 0,441632755 | 0,01863387 | 0,51104018 | 0,00545052 | 0,12706037 | 0,51937676 |
| ECAS_ExecFct | 0,390204378 | 0,04009048 | 0,41691562 | 0,0273046 | 0,14484651 | 0,46209405 |
| ECAS_Total | 0,71785161 | 1,70E-05 | 7,41E-01 | 6,52E-06 | 3,54E-01 | 0,06458835 |

**Supplementary Table 2:**
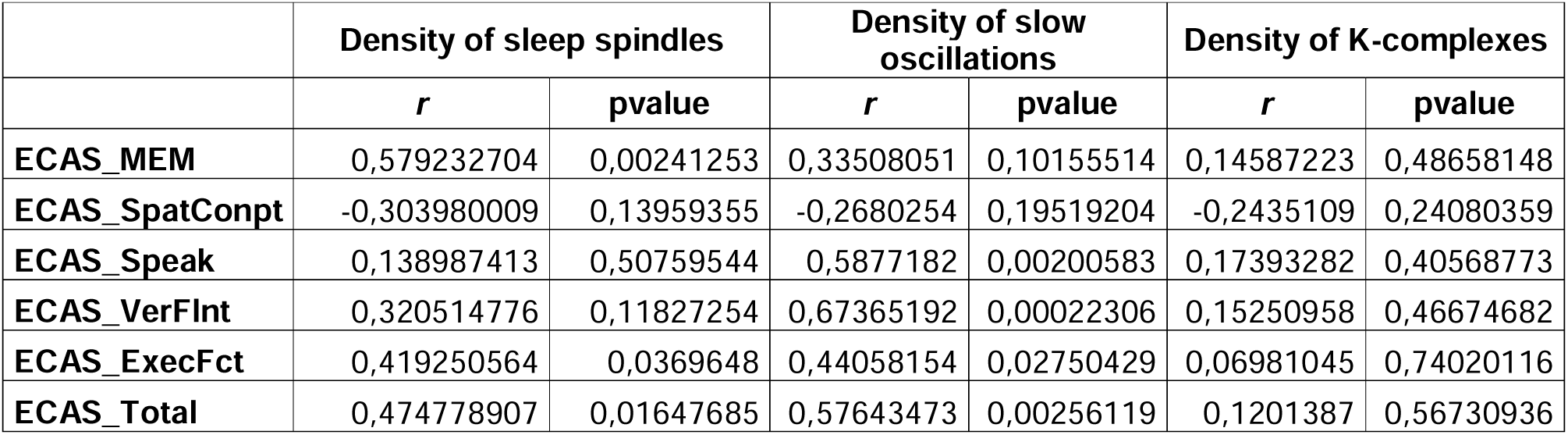
correlation between sleep microarchitecture and clinical parameters in presymptomatic gene carriers. Spearman r and adjusted p-values are indicated for each comparison.

## References

1. Goutman SA, Hardiman O, Al-Chalabi A, et al. Emerging insights into the complex genetics and pathophysiology of amyotrophic lateral sclerosis. Lancet Neurol. Mar 22 2022;doi:10.1016/S1474-4422(21)00414-2

2. Goutman SA, Hardiman O, Al-Chalabi A, et al. Recent advances in the diagnosis and prognosis of amyotrophic lateral sclerosis. Lancet Neurol. Mar 22 2022;doi:10.1016/S1474-4422(21)00465-8

3. Weydt P, Oeckl P, Huss A, et al. Neurofilament levels as biomarkers in asymptomatic and symptomatic familial amyotrophic lateral sclerosis. Ann Neurol. Jan 2016;79(1):152–8. doi:10.1002/ana.24552

4. Benatar M, Granit V, Andersen PM, et al. Mild motor impairment as prodromal state in amyotrophic lateral sclerosis: a new diagnostic entity. Brain. Oct 21 2022;145(10):3500–3508. doi:10.1093/brain/awac185

5. Benatar M, Wuu J, Huey ED, et al. The Miami Framework for ALS and related neurodegenerative disorders: an integrated view of phenotype and biology. Nat Rev Neurol. Jun 2024;20(6):364–376. doi:10.1038/s41582-024-00961-z

6. Diekmann K, Kuzma-Kozakiewicz M, Piotrkiewicz M, et al. Impact of comorbidities and co-medication on disease onset and progression in a large German ALS patient group. J Neurol. Jul 2020;267(7):2130–2141. doi:10.1007/s00415-020-09799-z

7. Janse van Mantgem MR, van Eijk RPA, van der Burgh HK, et al. Prognostic value of weight loss in patients with amyotrophic lateral sclerosis: a population-based study. J Neurol Neurosurg Psychiatry. Aug 2020;91(8):867–875. doi:10.1136/jnnp-2020-322909

8. Wei QQ, Ou R, Cao B, et al. Early weight instability is associated with cognitive decline and poor survival in amyotrophic lateral sclerosis. Brain Res Bull. Jun 2021;171:10–15. doi:10.1016/j.brainresbull.2021.02.022

9. Li JY, Sun XH, Cai ZY, et al. Correlation of weight and body composition with disease progression rate in patients with amyotrophic lateral sclerosis. Sci Rep. Aug 2 2022;12(1):13292. doi:10.1038/s41598-022-16229-9

10. Peter RS, Rosenbohm A, Dupuis L, et al. Life course body mass index and risk and prognosis of amyotrophic lateral sclerosis: results from the ALS registry Swabia. Article. Eur J Epidemiol. Oct 2017;32(10):901–908. doi:10.1007/s10654-017-0318-z

11. Abrahams S. Neuropsychological impairment in amyotrophic lateral sclerosis-frontotemporal spectrum disorder. Nat Rev Neurol. Nov 2023;19(11):655–667. doi:10.1038/s41582-023-00878-z

12. Lule DE, Muller HP, Finsel J, et al. Deficits in verbal fluency in presymptomatic C9orf72 mutation gene carriers-a developmental disorder. J Neurol Neurosurg Psychiatry. Nov 2020;91(11):1195–1200. doi:10.1136/jnnp-2020-323671

13. Guillot SJ, Lang C, Simonot M, et al. Early-onset sleep alterations found in patients with amyotrophic lateral sclerosis are ameliorated by orexin antagonist in mouse models. Sci Transl Med. Jan 29 2025;17(783):eadm7580. doi:10.1126/scitranslmed.adm7580

14. Fernandez LMJ, Luthi A. Sleep Spindles: Mechanisms and Functions. Physiol Rev. Apr 1 2020;100(2):805–868. doi:10.1152/physrev.00042.2018

15. Brodt S, Inostroza M, Niethard N, Born J. Sleep-A brain-state serving systems memory consolidation. Neuron. Apr 5 2023;111(7):1050–1075. doi:10.1016/j.neuron.2023.03.005

16. Dupuis L, de Tapia M, Rene F, et al. Differential screening of mutated SOD1 transgenic mice reveals early up-regulation of a fast axonal transport component in spinal cord motor neurons. Neurobiology of Disease. Aug 2000;7(4):274–285. doi:10.1006/nbdi.2000.0292

17. Dupuis L, Oudart H, Rene F, Gonzalez de Aguilar JL, Loeffler JP. Evidence for defective energy homeostasis in amyotrophic lateral sclerosis: benefit of a high-energy diet in a transgenic mouse model. Proc Natl Acad Sci U S A. Jul 27 2004;101(30):11159–64.

18. Scekic-Zahirovic J, Sendscheid O, El Oussini H, et al. Toxic gain of function from mutant FUS protein is crucial to trigger cell autonomous motor neuron loss. EMBO J. Mar 7 2016;35(10):1077–1097. doi:10.15252/embj.201592559

19. Scekic-Zahirovic J, El Oussini H, Mersmann S, et al. Motor neuron intrinsic and extrinsic mechanisms contribute to the pathogenesis of FUS-associated amyotrophic lateral sclerosis. Article. Acta Neuropathol (Berl). Jun 2017;133(6):887–906. doi:10.1007/s00401-017-1687-9

20. Arnold ES, Ling SC, Huelga SC, et al. ALS-linked TDP-43 mutations produce aberrant RNA splicing and adult-onset motor neuron disease without aggregation or loss of nuclear TDP-43. Proc Natl Acad Sci U S A. Feb 4 2013;doi:1222809110

21. Zhang Y, Ren R, Yang L, et al. Sleep in Alzheimer’s disease: a systematic review and meta-analysis of polysomnographic findings. Transl Psychiatry. Apr 1 2022;12(1):136. doi:10.1038/s41398-022-01897-y

22. D’Rozario AL, Chapman JL, Phillips CL, et al. Objective measurement of sleep in mild cognitive impairment: A systematic review and meta-analysis. Sleep Med Rev. Aug 2020;52:101308. doi:10.1016/j.smrv.2020.101308

23. Weng YY, Lei X, Yu J. Sleep spindle abnormalities related to Alzheimer’s disease: a systematic mini-review. Sleep Med. Nov 2020;75:37–44. doi:10.1016/j.sleep.2020.07.044

24. Villamar-Flores CI, Rodriguez-Violante M, Abundes-Corona A, et al. Association between alterations in sleep spindles and cognitive decline in persons with Parkinson’s disease. Neuroscience letters. Nov 1 2024;842:138006. doi:10.1016/j.neulet.2024.138006

25. Latreille V, Carrier J, Gaudet-Fex B, et al. Electroencephalographic prodromal markers of dementia across conscious states in Parkinson’s disease. Brain. Apr 2016;139(Pt 4):1189–99. doi:10.1093/brain/aww018

26. Christensen JA, Nikolic M, Warby SC, et al. Sleep spindle alterations in patients with Parkinson’s disease. Front Hum Neurosci. 2015;9:233. doi:10.3389/fnhum.2015.00233

27. Bender AC, Jaleel A, Pellerin KR, et al. Altered Sleep Microarchitecture and Cognitive Impairment in Patients With Temporal Lobe Epilepsy. Neurology. Dec 4 2023;101(23):e2376–e2387. doi:10.1212/WNL.0000000000207942

28. Mayeli A, Sanguineti C, Ferrarelli F. Recent Evidence of Non-Rapid Eye Movement Sleep Oscillation Abnormalities in Psychiatric Disorders. Curr Psychiatry Rep. Oct 14 2024;doi:10.1007/s11920-024-01544-x

29. Denis D, Baran B, Mylonas D, et al. Sleep oscillations and their relations with sleep-dependent memory consolidation in early course psychosis and first-degree relatives. Schizophr Res. Dec 2024;274:473–485. doi:10.1016/j.schres.2024.10.026

30. Ferrarelli F. Sleep spindles as neurophysiological biomarkers of schizophrenia. Eur J Neurosci. Apr 2024;59(8):1907–1917. doi:10.1111/ejn.16178

31. Peters KR, Ray LB, Fogel S, Smith V, Smith CT. Age differences in the variability and distribution of sleep spindle and rapid eye movement densities. PLoS One. 2014;9(3):e91047. doi:10.1371/journal.pone.0091047

32. Ameen MS, Petzka M, Peigneux P, Hoedlmoser K. Post-training sleep modulates motor adaptation and task-related beta oscillations. J Sleep Res. Aug 2024;33(4):e14082. doi:10.1111/jsr.14082

33. Hahn MA, Bothe K, Heib D, Schabus M, Helfrich RF, Hoedlmoser K. Slow oscillation-spindle coupling strength predicts real-life gross-motor learning in adolescents and adults. eLife. Feb 18 2022;11 doi:10.7554/eLife.66761

34. Boutin A, Doyon J. A sleep spindle framework for motor memory consolidation. Philos Trans R Soc Lond B Biol Sci. May 25 2020;375(1799):20190232. doi:10.1098/rstb.2019.0232

35. Kam K, Pettibone WD, Shim K, Chen RK, Varga AW. Dynamics of sleep spindles and coupling to slow oscillations following motor learning in adult mice. Neurobiol Learn Mem. Dec 2019;166:107100. doi:10.1016/j.nlm.2019.107100

36. Laventure S, Fogel S, Lungu O, et al. NREM2 and Sleep Spindles Are Instrumental to the Consolidation of Motor Sequence Memories. PLoS biology. Mar 2016;14(3):e1002429. doi:10.1371/journal.pbio.1002429

37. Wiesenfarth M, Huppertz HJ, Dorst J, et al. Structural and microstructural neuroimaging signature of C9orf72-associated ALS: A multiparametric MRI study. Neuroimage Clin. 2023;39:103505. doi:10.1016/j.nicl.2023.103505

38. Nigri A, Umberto M, Stanziano M, et al. C9orf72 ALS mutation carriers show extensive cortical and subcortical damage compared to matched wild-type ALS patients. Neuroimage Clin. 2023;38:103400. doi:10.1016/j.nicl.2023.103400

39. Bonham LW, Geier EG, Sirkis DW, et al. Radiogenomics of C9orf72 Expansion Carriers Reveals Global Transposable Element Derepression and Enables Prediction of Thalamic Atrophy and Clinical Impairment. J Neurosci. Jan 11 2023;43(2):333–345. doi:10.1523/JNEUROSCI.1448-22.2022

40. Chipika RH, Siah WF, Shing SLH, et al. MRI data confirm the selective involvement of thalamic and amygdalar nuclei in amyotrophic lateral sclerosis and primary lateral sclerosis. Data Brief. Oct 2020;32:106246. doi:10.1016/j.dib.2020.106246

41. Schonecker S, Neuhofer C, Otto M, et al. Atrophy in the Thalamus But Not Cerebellum Is Specific for C9orf72 FTD and ALS Patients - An Atlas-Based Volumetric MRI Study. Front Aging Neurosci. 2018;10:45. doi:10.3389/fnagi.2018.00045

42. Westeneng HJ, Walhout R, Straathof M, et al. Widespread structural brain involvement in ALS is not limited to the C9orf72 repeat expansion. J Neurol Neurosurg Psychiatry. Dec 2016;87(12):1354–1360. doi:10.1136/jnnp-2016-313959

43. Zhang JQ, Ji B, Zhou CY, et al. Differential Impairment of Thalamocortical Structural Connectivity in Amyotrophic Lateral Sclerosis. CNS Neurosci Ther. Feb 2017;23(2):155–161. doi:10.1111/cns.12658

44. Lee SE, Khazenzon AM, Trujillo AJ, et al. Altered network connectivity in frontotemporal dementia with C9orf72 hexanucleotide repeat expansion. Brain. Nov 2014;137(Pt 11):3047–60. doi:10.1093/brain/awu248

45. Shoukry RS, Waugh R, Bartlett D, Raitcheva D, Floeter MK. Longitudinal changes in resting state networks in early presymptomatic carriers of C9orf72 expansions. Neuroimage Clin. 2020;28:102354. doi:10.1016/j.nicl.2020.102354

46. Lajoie I, Canadian ALSNC, Kalra S, Dadar M. Regional Cerebral Atrophy Contributes to Personalized Survival Prediction in Amyotrophic Lateral Sclerosis: A Multicentre, Machine Learning, Deformation-Based Morphometry Study. Ann Neurol. Feb 22 2025;doi:10.1002/ana.27196

47. van Veenhuijzen K, Tan HHG, Nitert AD, et al. Longitudinal Magnetic Resonance Imaging in Asymptomatic C9orf72 Mutation Carriers Distinguishes Phenoconverters to Amyotrophic Lateral Sclerosis or Amyotrophic Lateral Sclerosis With Frontotemporal Dementia. Ann Neurol. Feb 2025;97(2):281–295. doi:10.1002/ana.27116

48. Mohammadi S, Ghaderi S, Mohammadi M, et al. Thalamic Alterations in Motor Neuron Diseases: A Systematic Review of MRI Findings. J Integr Neurosci. Apr 10 2024;23(4):77. doi:10.31083/j.jin2304077

49. Hinault T, Segobin S, Benbrika S, et al. Longitudinal grey matter and metabolic contributions to cognitive changes in amyotrophic lateral sclerosis. Brain Commun. 2022;4(5):fcac228. doi:10.1093/braincomms/fcac228

50. Machts J, Loewe K, Kaufmann J, et al. Basal ganglia pathology in ALS is associated with neuropsychological deficits. Neurology. Oct 13 2015;85(15):1301–9. doi:10.1212/WNL.0000000000002017

51. Tu S, Menke RAL, Talbot K, Kiernan MC, Turner MR. Regional thalamic MRI as a marker of widespread cortical pathology and progressive frontotemporal involvement in amyotrophic lateral sclerosis. J Neurol Neurosurg Psychiatry. Dec 2018;89(12):1250–1258. doi:10.1136/jnnp-2018-318625

52. Kolaj M, Zhang L, Hermes ML, Renaud LP. Intrinsic properties and neuropharmacology of midline paraventricular thalamic nucleus neurons. Front Behav Neurosci. 2014;8:132. doi:10.3389/fnbeh.2014.00132

53. Govindaiah G, Cox CL. Modulation of thalamic neuron excitability by orexins. Neuropharmacology. Sep 2006;51(3):414–25. doi:10.1016/j.neuropharm.2006.03.030

54. Bolborea M, Vercruysse P, Daria T, et al. Loss of hypothalamic MCH decreases food intake in amyotrophic lateral sclerosis. Acta Neuropathol. Jun 2023;145(6):773–791. doi:10.1007/s00401-023-02569-x

55. Vercruysse P, Vieau D, Blum D, Petersen A, Dupuis L. Hypothalamic Alterations in Neurodegenerative Diseases and Their Relation to Abnormal Energy Metabolism. Review. Front Mol Neurosci. Jan 2018;11:16. 2. doi:10.3389/fnmol.2018.00002

56. Gabery S, Ahmed RM, Caga J, Kiernan MC, Halliday GM, Petersen A. Loss of the metabolism and sleep regulating neuronal populations expressing orexin and oxytocin in the hypothalamus in amyotrophic lateral sclerosis. Neuropathology and applied neurobiology. Mar 23 2021;47(7):979–989. doi:10.1111/nan.12709

57. Inayat S, Qandeel, Nazariahangarkolaee M, et al. Low acetylcholine during early sleep is important for motor memory consolidation. Sleep. Jun 15 2020;43(6) doi:10.1093/sleep/zsz297

58. Gott JA, Stucker S, Kanske P, Haaker J, Dresler M. Acetylcholine and metacognition during sleep. Conscious Cogn. Jan 2024;117:103608. doi:10.1016/j.concog.2023.103608

59. Herrera CG, Tarokh L. A Thalamocortical Perspective on Sleep Spindle Alterations in Neurodevelopmental Disorders. Curr Sleep Med Rep. 2024;10(2):103–118. doi:10.1007/s40675-024-00284-x

60. Brunet A, Stuart-Lopez G, Burg T, Scekic-Zahirovic J, Rouaux C. Cortical Circuit Dysfunction as a Potential Driver of Amyotrophic Lateral Sclerosis. Front Neurosci. 2020;14:363. doi:10.3389/fnins.2020.00363

61. Luthi A, Nedergaard M. Anything but small: Microarousals stand at the crossroad between noradrenaline signaling and key sleep functions. Neuron. Feb 19 2025;113(4):509–523. doi:10.1016/j.neuron.2024.12.009

62. Osorio-Forero A, Cardis R, Vantomme G, et al. Noradrenergic circuit control of non-REM sleep substates. Curr Biol. Nov 22 2021;31(22):5009–5023 e7. doi:10.1016/j.cub.2021.09.041

63. Swift KM, Gross BA, Frazer MA, et al. Abnormal Locus Coeruleus Sleep Activity Alters Sleep Signatures of Memory Consolidation and Impairs Place Cell Stability and Spatial Memory. Curr Biol. Nov 19 2018;28(22):3599–3609 e4. doi:10.1016/j.cub.2018.09.054

64. Scekic-Zahirovic J, Benetton C, Brunet A, et al. Cortical hyperexcitability in mouse models and patients with amyotrophic lateral sclerosis is linked to noradrenaline deficiency. Sci Transl Med. Mar 13 2024;16(738):eadg3665. doi:10.1126/scitranslmed.adg3665

65. Walker NA, Sunderram J, Zhang P, Lu SE, Scharf MT. Clinical utility of the Epworth sleepiness scale. Sleep Breath. Dec 2020;24(4):1759–1765. doi:10.1007/s11325-020-02015-2

66. Buysse DJ, Reynolds CF, 3rd, Monk TH, Berman SR, Kupfer DJ. The Pittsburgh Sleep Quality Index: a new instrument for psychiatric practice and research. Psychiatry Res. May 1989;28(2):193–213. doi:10.1016/0165-1781(89)90047-4

67. Lule D, Burkhardt C, Abdulla S, et al. The Edinburgh Cognitive and Behavioural Amyotrophic Lateral Sclerosis Screen: a cross-sectional comparison of established screening tools in a German-Swiss population. Amyotrophic lateral sclerosis & frontotemporal degeneration. Mar 2015;16(1-2):16–23. doi:10.3109/21678421.2014.959451

68. Abrahams S, Newton J, Niven E, Foley J, Bak TH. Screening for cognition and behaviour changes in ALS. Amyotrophic lateral sclerosis & frontotemporal degeneration. Mar 2014;15(1-2):9–14. doi:10.3109/21678421.2013.805784

69. Loose M, Burkhardt C, Aho-Ozhan H, et al. Age and education-matched cut-off scores for the revised German/Swiss-German version of ECAS. Amyotrophic lateral sclerosis & frontotemporal degeneration. Jul-Aug 2016;17(5-6):374–6. doi:10.3109/21678421.2016.1162814

70. Berry RB, Budhiraja R, Gottlieb DJ, et al. Rules for scoring respiratory events in sleep: update of the 2007 AASM Manual for the Scoring of Sleep and Associated Events. Deliberations of the Sleep Apnea Definitions Task Force of the American Academy of Sleep Medicine. J Clin Sleep Med. Oct 15 2012;8(5):597–619. doi:10.5664/jcsm.2172

71. Silber MH, Ancoli-Israel S, Bonnet MH, et al. The visual scoring of sleep in adults. J Clin Sleep Med. Mar 15 2007;3(2):121–31.

72. Ablin P, Cardoso JF, Gramfort A. Faster Independent Component Analysis by Preconditioning With Hessian Approximations. IEEE Transactions on Signal Processing. 2018;66(15):4040–4049. doi:10.1109/tsp.2018.2844203

73. Dammers J, Schiek M, Boers F, et al. Integration of amplitude and phase statistics for complete artifact removal in independent components of neuromagnetic recordings. IEEE Trans Biomed Eng. Oct 2008;55(10):2353–62. doi:10.1109/TBME.2008.926677

74. Dharmaprani D, Nguyen HK, Lewis TW, DeLosAngeles D, Willoughby JO, Pope KJ. A comparison of independent component analysis algorithms and measures to discriminate between EEG and artifact components. Annu Int Conf IEEE Eng Med Biol Soc. Aug 2016;2016:825–828. doi:10.1109/EMBC.2016.7590828

75. Viola FC, Thorne J, Edmonds B, Schneider T, Eichele T, Debener S. Semi-automatic identification of independent components representing EEG artifact. Clin Neurophysiol. May 2009;120(5):868–77. doi:10.1016/j.clinph.2009.01.015

76. Winkler I, Debener S, Muller KR, Tangermann M. On the influence of high-pass filtering on ICA-based artifact reduction in EEG-ERP. Annu Int Conf IEEE Eng Med Biol Soc. 2015;2015:4101–5. doi:10.1109/EMBC.2015.7319296

77. Lacourse K, Delfrate J, Beaudry J, Peppard P, Warby SC. A sleep spindle detection algorithm that emulates human expert spindle scoring. J Neurosci Methods. Mar 15 2019;316:3–11. doi:10.1016/j.jneumeth.2018.08.014

78. Massimini M, Huber R, Ferrarelli F, Hill S, Tononi G. The sleep slow oscillation as a traveling wave. J Neurosci. Aug 4 2004;24(31):6862–70. doi:10.1523/JNEUROSCI.1318-04.2004

79. Carrier J, Viens I, Poirier G, et al. Sleep slow wave changes during the middle years of life. Eur J Neurosci. Feb 2011;33(4):758–66. doi:10.1111/j.1460-9568.2010.07543.x

80. Virtanen P, Gommers R, Oliphant TE, et al. SciPy 1.0: fundamental algorithms for scientific computing in Python. Nature methods. Mar 2020;17(3):261–272. doi:10.1038/s41592-019-0686-2

81. Vallat R, Walker MP. An open-source, high-performance tool for automated sleep staging. eLife. Oct 14 2021;10 doi:10.7554/eLife.70092

82. Vallat R, Walker MP. An open-source, high-performance tool for automated sleep staging. Elife. Oct 14 2021;10:e70092. doi:10.7554/eLife.70092

83. American Academy of Sleep Medicine. The AASM Manual for the Scoring of Sleep and Associated Events, Scoring Manual Version 2.6. Darien, IL, USA: AASM; 2020.

84. Guillot SJ. Sleep spindles detector. Version 2023. https://github.com/sjg2203/SSp_Detector

85. Guillot SJ. SlµArch. Version 2024. https://github.com/sjg2203/SluArch

86. Faul F, Erdfelder E, Buchner A, Lang AG. Statistical power analyses using G*Power 3.1: tests for correlation and regression analyses. Behav Res Methods. Nov 2009;41(4):1149–60. doi:10.3758/BRM.41.4.1149

87. Faul F, Erdfelder E, Lang AG, Buchner A. G*Power 3: a flexible statistical power analysis program for the social, behavioral, and biomedical sciences. Behav Res Methods. May 2007;39(2):175–91. doi:10.3758/bf03193146

88. SHAPIRO SS, WILK MB. An analysis of variance test for normality (complete samples). Biometrika. 1965;52(3-4):591–611. doi:10.1093/biomet/52.3-4.591

89. Bartlett MS. Properties of sufficiency and statistical tests. . Proceedings of the Royal Society of London Series A-Mathematical and Physical Sciences 1937;160:268–282.

90. Vallat R. Pingouin: statistics in Python. Journal of Open Source Software. 2018;3:1026.

91. Zimmerman DW. A note on preliminary tests of equality of variances. Br J Math Stat Psychol. May 2004;57(Pt 1):173–81. doi:10.1348/000711004849222

92. Rouder JN, Speckman PL, Sun D, Morey RD, Iverson G. Bayesian t tests for accepting and rejecting the null hypothesis. Psychon Bull Rev. Apr 2009;16(2):225–37. doi:10.3758/PBR.16.2.225

93. Seabold S, Perktold J. Statsmodels: Econometric and Statistical Modeling with Python. 2010:25080.

94. Terpilowski M. scikit-posthocs: Pairwise multiple comparison tests in Python. Journal of Open Source Software. 2019;4:1169.

95. Waskom M. seaborn: statistical data visualization. Journal of Open Source Software 2021;6:3021.

